# Childhood sleep trajectories, hypomanic symptoms, and cognitive functioning in early adulthood: a longitudinal cohort study

**DOI:** 10.64898/2026.11.19.26368486

**Authors:** Buse Beril Durdurak, Steven Marwaha, Paul R.A. Stokes, Isabel Morales-Muñoz

## Abstract

**Background:** Sleep and circadian rhythm disturbances are implicated in bipolar disorder (BD), yet it remains unclear whether developmental trajectories of sleep and circadian rhythms across childhood confer risk for hypomanic symptoms (HS) and cognitive dysfunction. Here, we examined whether distinct sleep duration and sleep timing trajectories across childhood are associated with potential HS and cognitive functioning in early adulthood.

**Methods:** Using data from the Avon Longitudinal Study of Parents and Children (ALSPAC), parent-reported night-time sleep duration and mid-point of sleep (MPS) from 6 months to 11 years were modelled using latent class growth analysis. During the ALSPAC study, HS were assessed at ages 21–23 using the Hypomania Checklist (HCL), and cognitive functioning (working memory and response inhibition) was assessed at age 24.

**Results:** Four sleep duration and three MPS trajectories were identified. Logistic regression analyses showed that persistently average shorter (OR=2.87, 95% CI=1.31–6.32) and persistently longer sleep (OR=2.81, 95% CI=1.08–7.30) trajectories were associated with increased risk of potential HS, while persistently earlier MPS was associated with lower odds (OR=0.23, 95% CI=0.11–0.49). Multiple linear regression models showed that experiencing HS in combination with persistently average shorter sleep was associated with poorer response inhibition (B=36.55, 95% CI=8.33–64.78, p=0.011), whereas HS in combination with persistently longer sleep was associated with better response inhibition (B=−125.21, 95% CI= −193.56 to −56.86, p<0.001). No interactions were found for working memory.

**Conclusions:** Persistently average shorter and persistently longer sleep duration trajectories were associated with increased risks of experiencing HS, as measured by the HCL, while earlier circadian timing were associated with reduced risks. Differences in early-life sleep regulation may be associated with subsequent risk of experiencing HS during early adulthood and may differentially shape subsequent response inhibition performance.

## Introduction

Sleep and circadian rhythm disturbances (SCRD) are core clinical features of bipolar disorder (BD) and are increasingly recognised not only as correlates of illness state, but also as potential markers of vulnerability [1,2]. Disturbances in sleep duration, timing, and rhythmicity occur across illness phases of BD and are implicated in episode onset, recurrence, and illness course [2–4]. Longitudinal studies further suggest that sleep disturbances often precede the onset of BD, potentially functioning as early risk markers [4–7]. A systematic review and meta-analysis found that eveningness and social rhythm dysregulation are elevated in individuals at risk for BD, with pre-existing SCRD conferring approximately 40% increased risk of later onset of BD [2]. Findings regarding sleep duration, however, are less consistent, with both shorter sleep [8] and longer sleep duration [2,9] implicated in the risk of developing BD. [9]

Despite the high prevalence of SCRD across early phases of BD, most prior research has assessed sleep at single timepoints or focused only on specific subgroups at risk of BD and/or within a peak age range for onset of BD [2]. Characterising developmental trajectories rather than isolated measures can help disentangle transient from persistent sleep problems and identify subgroups with stable deviations from typical developmental patterns [10]. One way to identify the earliest clinical manifestations of BD are to study hypomanic symptoms (HS) [11], a common feature of BD in youth which may be associated with subsequent manic episodes [12]. To date, however, no population-based study has examined whether sleep trajectories from infancy to late childhood may be associated with HS in young adulthood.

SCRD are associated with poorer working memory, attention, and response inhibition both in healthy adults [13–15] and healthy young people [16,17]. A number of studies have suggested a significant association between sleep disturbance and cognitive impairment in mood disorders[18]. Within BD, cognitive impairments, such as poorer executive functioning and working memory are prevalent, persist beyond mood episodes into euthymia, and contribute to functional disability [19–21]. However, there is marked heterogeneity in cognitive impairments across people with BD [22], making it important to identify factors that may contribute to this variation. Crucially, sleep dysregulation may contribute to, or exacerbate, cognitive impairment in BD [18,23]. For example, an actigraphy study showed that people with BD with abnormal sleep displayed slower processing speed and attentional deficits compared to healthy participants and BD participants with normal sleep [24]. Similarly, another study found that individuals with BD reporting any sleep disturbance performed worse on processing speed and inhibition than those without sleep disturbance [25]. However, it remains unclear whether developmental patterns of sleep are associated with later cognitive impairments among young people who may be at risk of developing HS. This is important because sleep is observable from early life and potentially modifiable.

Building on this rationale, we firstly sought to characterise the developmental trajectories of night-time sleep duration and sleep timing in children assessed from 6 months to 11 years of age, and to evaluate their prospective associations with HS as measured using the Hypomania Checklist Questionnaire (HCL-32) assessed at the age of 21–23 years. Secondly, we sought to examine whether the combination of certain sleep duration and timing trajectories with the experience of HS in young adulthood was associated with poorer working memory and response inhibition performance assessed at age 24. We specifically sought to examine whether childhood sleep trajectories modified the association between HS at ages 21–23 and two measures of cognitive functioning, working memory and response inhibition, at age 24. We focused on working memory and response inhibition because they capture complementary aspects of executive control, and previous studies have linked sleep disruption to performance in both of these cognitive domains [14–18].

## Methods

### Participants

This study used data from the Avon Longitudinal Study of Parents and Children (ALSPAC), an ongoing longitudinal UK birth cohort study designed to investigate the factors associated with the development, health, and disease during childhood and beyond [26–28]. Pregnant women resident in Avon, UK, with expected dates of delivery between 1 April 1991 and 31 December 1992 were invited to take part in the study. Of 20,248 eligible pregnancies, 14,541 were initially enrolled, resulting in 13,988 children alive at one year of age [26,27]. Additional participants were enrolled from age seven onwards (see Supplement for further details and Figure S1 for the sample flow chart).

Ethical approval for the ALSPAC study was obtained from the ALSPAC Ethics and Law Committee and the Local Research Ethics Committees. The age-24 clinic assessments used in our analysis were approved by the National Research Ethics Service Committee South West–Frenchay (14/SW/1173). Informed consent for the use of all data collected was obtained by ALSPAC from participants following the recommendations of the ALSPAC Ethics and Law Committee at the time.

Data for the present study were accessed through ALSPAC’s managed data-access process, following approval of the research proposal by the ALSPAC Executive Committee and in accordance with the relevant data-access agreements and governance requirements. Further details of the consent procedures are provided in the Supplementary Methods.

### Measures

All measures below were extracted from ALSPAC assessments taken during childhood, adolescence and early adulthood.

#### Night-time sleep duration across infancy and childhood

Childhood night-time sleep duration was estimated via parent report data. Parents were asked what time their child normally went to sleep at night and awoke in the morning. Then, the difference between these two timepoints was used to derive night-time sleep duration at each timepoint. This information was collected at 6, 18, and 30 months, and at 3.5, 4.5, 5.5, 6.5, 9 and 11 years of age.

#### Sleep timing (i.e., Mid-point of Sleep) across infancy and childhood

Mid-point of Sleep (MPS) was measured at 6, 18, and 30 months, and at 3.5, 4.5, 5.5, 6.5, 9 and 11 years of age, and was calculated as the middle timepoint between bedtime and wakeup time. MPS was used as an indirect marker of sleep timing. MPS correlates with dim-light melatonin onset across infancy, childhood and adulthood and outperforms bedtime alone as a circadian proxy [29,30].

#### Measures of hypomanic symptoms in young people at 21-23 years

Participants completed the HCL-32 [31], a self-report measure of lifetime HS (which is not clinically verified) comprising of 32 items, when they were 21-23 years old. Consistent with previous work, we constructed a dichotomous HS variable; a) those with a symptom score of 14 or more (out of 32) were classed as experiencing HS if they also reported, b) at least one incident of “negative consequences” or of “negative plus positive consequences,” as a result of these symptoms, c) that mood changes caused a reaction in people close to the participant and d) that symptoms lasted for “4 days” or more.

#### Cognitive Measures at 24 years

Working memory was assessed in the ALSPAC study using an n-back task [32], with d′ as the primary outcome; higher values indicate better discrimination (see Supplementary-Methods for further details). Response inhibition was assessed using a Stop Signal Task [33], with stop-signal reaction time (SSRT) as the primary outcome; lower values indicate better inhibition (see Supplementary-Methods for further details).

### Confounders

Covariates selected a priori included child’s sex at birth, child’s ethnicity, mother’s age at birth, gestational age, borderline personality disorder (BPD) traits at age 11 [34] and early family adversity, including socioeconomic disadvantage, maternal psychopathology, and adverse family conditions. BPD traits were included given previous evidence linking childhood sleep disturbances with borderline personality traits in adolescence, and adolescent borderline personality traits with subsequent hypomanic symptoms [35,36]. Rationale and measurement detail are provided in Supplementary-Methods.

### Statistical Analyses

Latent class growth analyses (LCGAs) were conducted in Mplus, version 8 [37], to identify distinct trajectories of night-time sleep duration and MPS from infancy to adolescence, analysed separately. Indicator variables included night-time sleep duration and MPS measured at 6, 18, and 30 months, and at ages 3.5, 4.5, 5.5, 6.5, 9, and 11 years. Several models were fit by increasing the number of classes [38]. Model fit was assessed using Bayesian information criterion, Lo-Mendell-Rubin likelihood ratio test, Vuong-Lo-Mendell-Rubin test, entropy value and proportion of individuals in each class [38]. In determining the optimal class solution, we prioritised model fit, adequate class sizes, distinctiveness, and clinical interpretability (see Supplementary-Methods). Missing data due to attrition were addressed using full information maximum likelihood estimation [39].

#### Primary Analyses

Using the classes identified by LCGA, we conducted two separate binary logistic regressions to explore the prospective associations of night-time sleep duration and MPS with HS at 21-23 years, one per sleep/chronotype domain. We conducted logistic regression analyses using SPSS, version 27 (IBM Corp). The derived latent classes from the LCGA were the predictors (with the class with larger sample as reference), and HS at 21-23 years was the outcome. We then tested unadjusted associations, as well as adjusted associations controlling for all confounders.

#### Secondary Analyses

To investigate whether the interaction between night-time sleep duration and MPS trajectory classes across childhood with experiencing HS at age 21-23 was associated with poorer working memory and response inhibition performance, we conducted multiple linear regression analyses. For each sleep trajectory previously associated with experiencing HS in the logistic regressions, separate binary variables were created contrasting the class of interest (coded 1) with the normative reference class (coded 0). For each trajectory, an interaction term was computed by multiplying the binary hypomanic symptom status with the binary sleep class variable. Hierarchical linear regression models were estimated separately for each sleep trajectory, with cognitive performance, working memory (i.e., n-back d′) and response inhibition (i.e., SSRT) modelled separately, as the outcome. Model 1 included hypomanic symptom status and sleep class as main effects. Model 2 added the interaction term to test moderation. Model 3 further adjusted for the same confounders as above.

#### Missing Data in Primary and Secondary Analyses

Among 15,645 participants in the original sample of ALSPAC, 13,951 participants were lost to follow-up at age 21-23 years. Therefore, to deal with missingness, we conducted weighted analysis using inverse probability to account for those lost to follow-up (See Supplementary-Methods). For the primary analyses, characteristics associated with attrition at 21-23 years old were being male, having a younger mother who had lower socioeconomic levels, and higher scores on FAI (see Supplementary-Table S3). Using the variables associated with selective dropout as the factors to predict missingness in our analysis sample, we fitted a logistic regression model (nonresponse vs response outcome) to determine weights for each individual using the inverse probability of response. For the secondary analyses, the same procedure was applied separately using the factors associated with attrition on the Stop Signal and N-back tasks (Supplementary-Tables S4–S5), yielding task-specific weights for the corresponding cognitive models.

## Results

Table 1 shows the frequencies and descriptive values of the variables of interest in this study.

**Table 1.** Descriptive values of sociodemographic variables, childhood sleep measures, hypomanic symptoms and cognitive outcomes in the ALSPAC Cohort^a^.

| **Variable** | **Mean (SD)** | **N (%)** |
|---|---|---|
| Child’s Sex |  |  |
| Female | - | 7348 (48.9) |
| Male | - | 7691 (51.1) |
| Ethnicity |  |  |
| White | - | 12062 (97.4) |
| Bangladeshi | - | 7 (0.1) |
| Black African | - | 11 (0.1) |
| Black Caribbean | - | 76 (0.6) |
| Black Other | - | 44 (0.4) |
| Chinese | - | 30 (0.2) |
| Indian | - | 54 (0.4) |
| Pakistani | - | 22 (0.2) |
| Other | - | 82 (0.7) |
| Maternal age at birth (in years) | 27.99 (4.96) | - |
| Gestational age (in weeks) | 38.36 (5.51) | - |
| Nighttime sleep duration (in hours) |  |  |
| 6 months | 10.80 (1.36) | - |
| 18 months | 11.33 (1.08) | - |
| 30 months | 11.24 (1.01) | - |
| 3.5 years | 11.27 (0.90) | - |
| 4.5 years | 11.39 (0.72) | - |
| 5.5 years | 11.28 (0.72) | - |
| 6.5 years | 10.99 (0.64) | - |
| 9 years | 10.43 (0.65) | - |
| 11 years | 10.05 (0.66) | - |
| Mid-Point of Sleep (in hh:mm) |  |  |
| 6 months | 1:32 (0:52) | - |
| 18 months | 1:28 (0:51) | - |
| 30 months | 1:26 (0:41) | - |
| 3.5 years | 1:23 (0:37) | - |
| 4.5 years | 1:26 (0:32) | - |
| 5.5 years | 1:31 (0:30) | - |
| 6.5 years | 1:54 (0:32) | - |
| 9 years | 2:24 (0:29) | - |
| 11 years | 2:45 (0:29) | - |
| FAI, total scores<sup>b</sup> | 4.38 (4.31) | - |
| BPD symptoms at 11 years, total score | 0.35 (0.86) | - |
| HCL-32 total symptom score at 21–23 years<sup>c</sup> | 15.14 (16.00) | - |
| Dichotomous HS variable at 21-23 years |  |  |
| Yes | - | 25 (1.5) |
| No | - | 1669 (98.5) |
| Stop-signal reaction time (SSRT), in milliseconds | 259.14 (52.87) | - |
| N-back d′ | 2.19 (0.91) | - |

Note: N= Total number of participants; BPD = Borderline Personality Disorder; SD, Standard Deviation.
<sup>a</sup> Unweighted descriptive values for the total sample.
<sup>b</sup> The total Family Adversity Index scores for 3 time-points (i.e., during pregnancy, age 2 years, and age 4 years) were summed.
<sup>c</sup> Participants were asked to consider a time when they were in a “high or hyper” state and endorse a number of statements about their emotions, thoughts, and behaviours at that time.

### Latent classes of night-time sleep duration and MPS

Table 2 shows the values of log-likelihood VLMR, ABIC, and number of other parameters for all models assessed. Overall, a four-class model offered the best model fit and theoretical explanation for latent classes of night-time sleep duration (See Supplementary-Methods for further detail). The four derived classes are shown in Figure 1: Class 1, persistently shorter sleep duration (n = 444 [3.1%]); Class 2, persistently longer sleep duration (n = 1,890 [13.4%]); Class 3, persistently average longer sleep duration (n = 7,962 [56.3%]); and Class 4, persistently average shorter sleep duration (n = 3,853 [27.2%]). The two average sleep-duration classes represented intermediate patterns, with the persistently average shorter class showing longer sleep duration than the persistently shorter class but shorter duration than the persistently average longer class, which in turn showed shorter duration than the persistently longer class.

**Figure 1.**
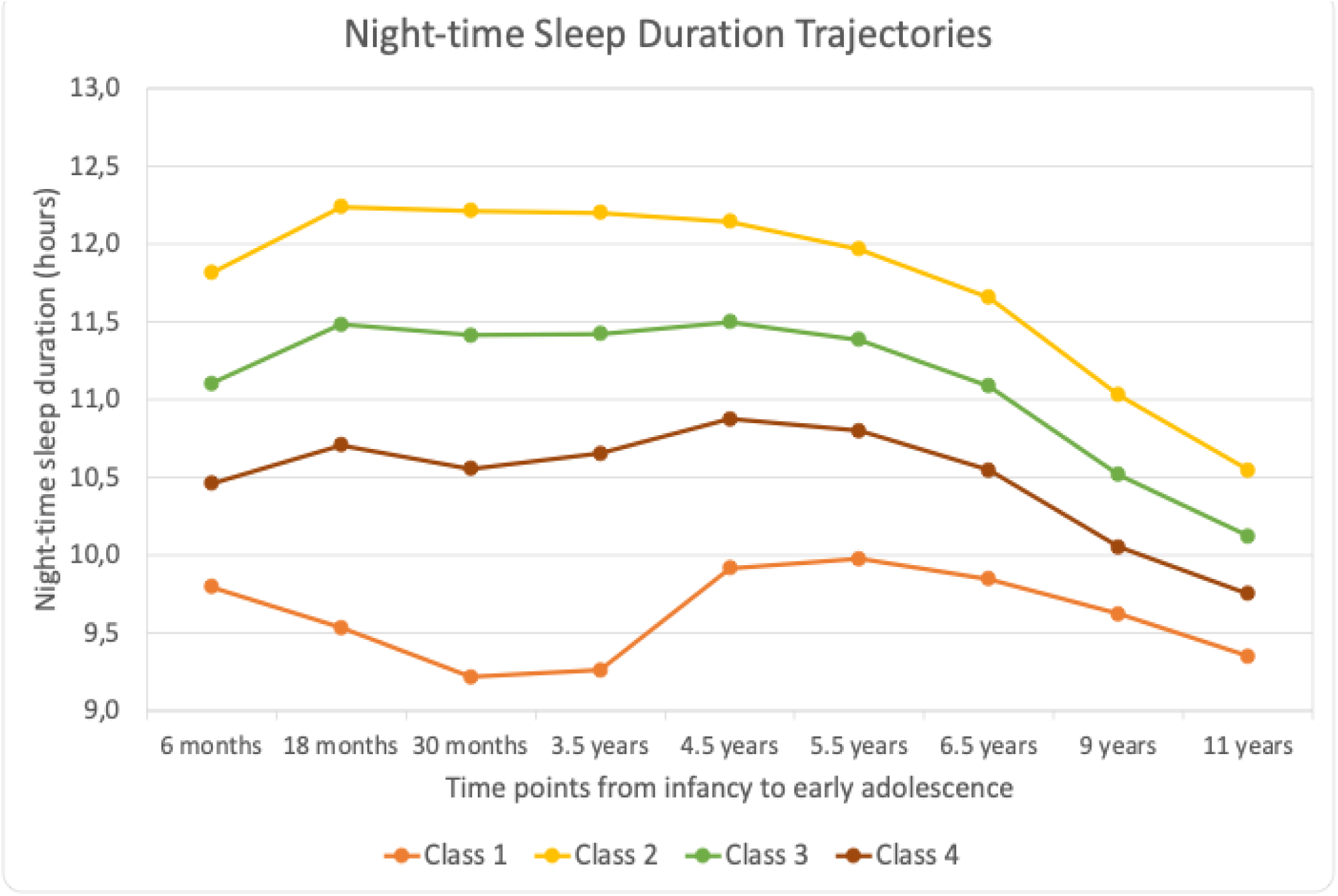
Four class model of nighttime sleep duration trajectories from 6 months to 11 years old. Growth trajectories of night-time sleep duration across infancy, childhood and early adolescence (from 6 months to 11 years). The latent class growth analyses detected a best model fit for 4 classes. X axis represents the 9 time points from infancy to early adolescence, and the Y axis represents the mean number of hours. Class 1 (orange line, 444 participants [3.1%]) represents individuals with persistently shorter sleep duration. Class 2 (yellow line, 1,890 participants [13.4%]) represents individuals with persistently longer sleep duration. Class 3 (green line, 7,962 participants [56.3%]) represents individuals with persistently average longer sleep duration across time points. Class 4 (brown line, 3,853 participants [27.2%]) represents individuals with persistently average shorter sleep duration.

**Table 2.**
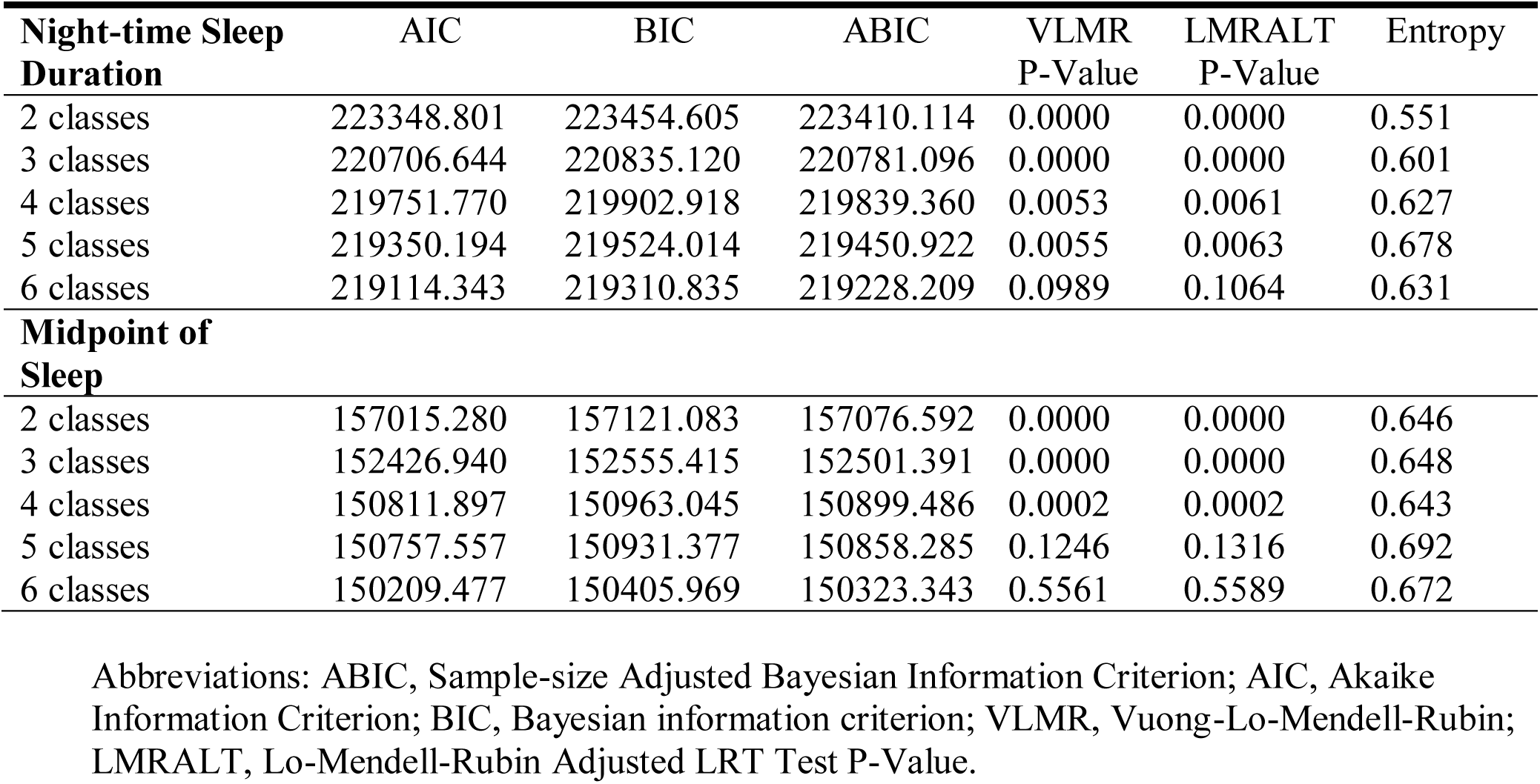
BIC, VLMR likelihood test p values, and entropy for classes 2–6 of the night-time sleep duration and midpoint of sleep variables.

For MPS, a 3-class model offered the best model fit and theoretical explanation (See Supplementary-Methods for further detail). The 3 derived classes are shown in Figure 2. Class 1 represented persistently earlier MPS (4,707 participants [33.3%]); class 2, persistently average earlier MPS (8,018 [56.7%]); and class 3, persistently later MPS (1,424 [10.1%]).

**Figure 2.**
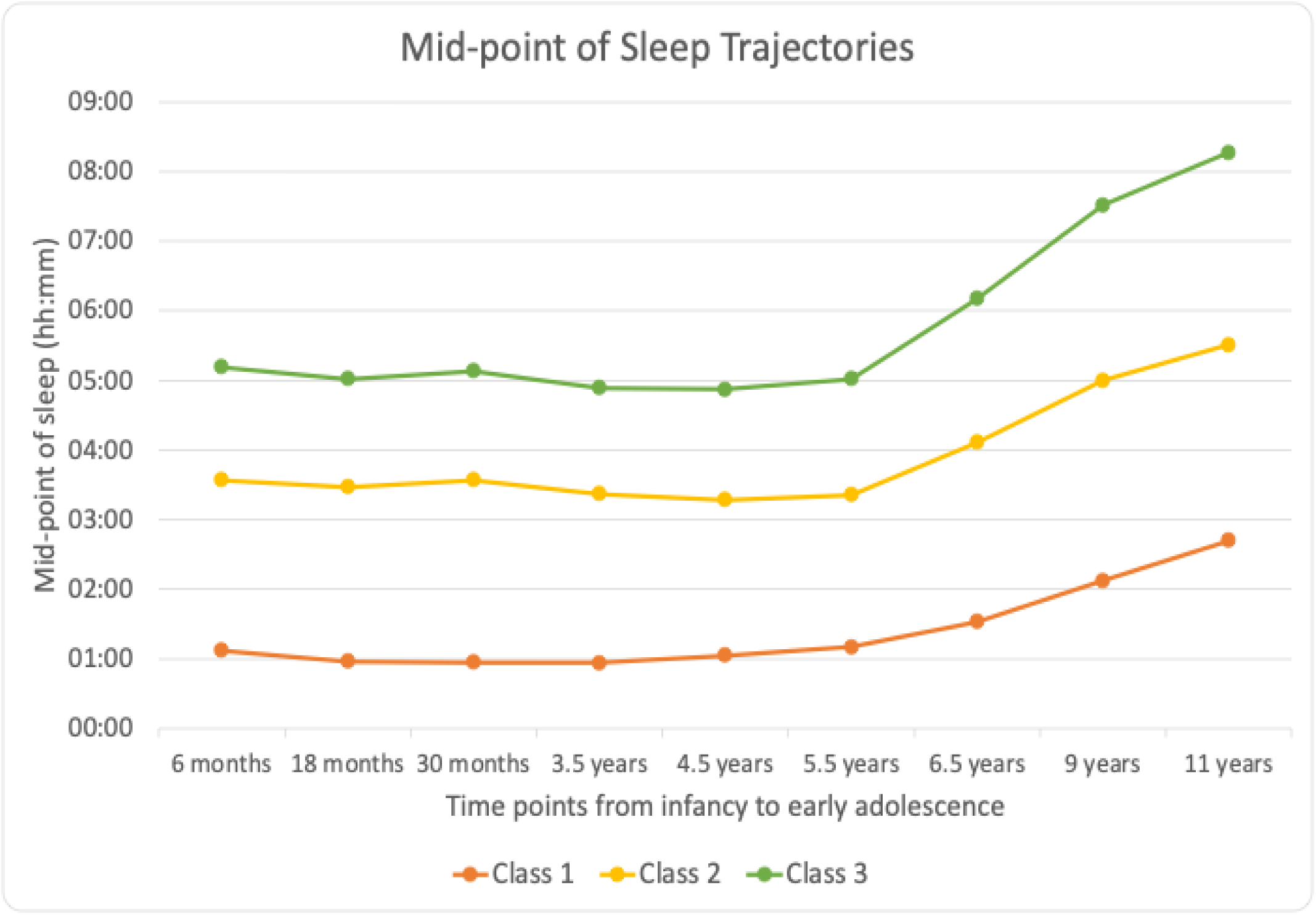
Three class model of Mid-Point of Sleep trajectories from 6 months to 11 years old. Growth trajectories of midpoint of sleep (MPS) across infancy, childhood and early adolescence (from 6 months to 11 years). The latent class growth analyses detected a best model fit for 3 classes. X axis represents the 9 time points from infancy to early adolescence, and the Y axis represents the mean clock time. Class 1 (orange line, 4707 participants [33.3%]) represents individuals with persistently earlier MPS across time points. Class 2 (yellow line, 8018 participants [56.7%]) represents individuals with persistently average earlier MPS. Class 3 (green line, 1424 participants [10.1%]) represents individuals with persistently later MPS.

### Night-time sleep duration classes and risk of experiencing hypomanic symptoms

The weighted adjusted logistic regression model (with persistently average longer sleep duration as the reference) showed that persistently average shorter class (OR=2.87; 95% CI=1.31-6.32; p=0.009) and persistently longer class (OR=2.81; 95% CI=1.08-7.30; p=0.035) were significantly associated with a higher risk of experiencing HS at the age of 21-23 (Table 3). None of the 16 participants in the persistently shorter sleep-duration class with available hypomanic symptom data met the criteria for hypomanic symptoms. Consequently, an odds ratio could not be reliably estimated for this class (Supplementary Methods S3.5 and Table S1).

**Table 3.** Associations of latent classes of night-time sleep duration and midpoint of sleep and risk of experiencing hypomanic symptoms at 21-23 years^a^.

|  | Unadjusted Model |  |  | Adjusted Model |  |  |
|---|---|---|---|---|---|---|
|  | OR | 95% CI | *P* Value | OR | 95% CI | *P* Value |
| **Night-time Sleep Duration** |  |  |  |  |  |  |
| Persistently Average Longer Class (Ref) | - | - | 0.011 | - | - | 0.045 |
| Persistently Average Shorter Class | 3.37 | 1.61-7.05 | 0.001 | 2.87 | 1.31-6.32 | 0.009 |
| Persistently Longer Class | 2.93 | 1.15-7.46 | 0.024 | 2.81 | 1.08-7.30 | 0.035 |
| Persistently Shorter Class | 0.00 | 0.00 | 0.998<sup>b</sup> | 0.00 | 0.00 | 0.998<sup>b</sup> |
| Child’s sex | - | - | - | 1.02 | 0.51-2.07 | 0.947 |
| Child’s ethnicity | - | - | - | 0.00 | 0.00 | 0.996 |
| Maternal Age at birth | - | - | - | 0.95 | 0.88-1.04 | 0.279 |
| Gestational Age | - | - | - | 1.25 | 1.00-1.55 | 0.047 |
| Family Adversity Index | - | - | - | 1.05 | 0.99-1.11 | 0.124 |
| BPD traits at 11 years | - | - | - | 1.39 | 1.12-1.73 | 0.003 |
| **Midpoint of Sleep** |  |  |  |  |  |  |
| Persistently Average Earlier Class (Ref) | - | - | <0.001 | - | - | <0.001 |
| Persistently Later Class | 0.00 | 0.00 | 0.993<sup>c</sup> | 0.00 | 0.00 | 0.993<sup>c</sup> |
| Persistently Earlier Class | 0.21 | 0.10-0.44 | <0.001 | 0.23 | 0.11-0.49 | <0.001 |
| Child’s sex | - | - | - | 0.82 | 0.47-1.44 | 0.488 |
| Child’s ethnicity | - | - | - | 0.00 | 0.00 | 0.996 |
| Maternal Age at birth | - | - | - | 0.94 | 0.88-1.00 | 0.043 |
| Gestational Age | - | - | - | 1.12 | 0.95-1.32 | 0.182 |
| Family Adversity Index | - | - | - | 1.09 | 1.05-1.14 | <0.001 |
| BPD traits at 11 years | - | - | - | 1.23 | 1.02-1.48 | 0.030 |

Note: BPD = Borderline Personality Disorder; OR = Odds Ratio.
<sup>a</sup> All analyses were weighted for sex, ethnicity, maternal age, maternal socioeconomic status, preterm delivery, birthweight and family adversity; Adjusted Model: associations adjusted for child’s sex, child’s ethnicity, maternal age at birth, gestational age, family adversity scores during pregnancy, at 2 years of age and 4 years of age, and BPD traits at 11 years.
<sup>b</sup> The persistently shorter sleep duration class contained no participants with hypomanic symptoms (complete separation); its odds ratio is not estimable and is reported as 0.00 (Supplementary Table S1).
<sup>c</sup> The persistently later MPS class contained only one participant with hypomanic symptoms (quasi-complete separation); its odds ratio is not reliably estimable and is reported as 0.00 (Supplementary Table S2).

### MPS classes and risk of experiencing hypomanic symptoms

The weighted adjusted logistic regression model (with persistently average earlier MPS as the reference) showed that persistently earlier MPS class (OR=0.23; 95% CI=0.11-0.49; p<0.001) was significantly associated with a lower risk of experiencing HS at the age of 21-23 (Table 3). Only one of the 150 participants in the persistently later MPS class with available hypomanic symptom data met the criteria for hypomanic symptoms. Consequently, the weighted odds ratio for this class could not be reliably estimated (Supplementary Methods S3.5 and Table S2).

### Sleep trajectories, hypomanic symptoms, and working memory

No significant interactions were observed between HS and childhood sleep duration or MPS trajectories with working memory at age 24 (Table 4). However, in the adjusted models, HS were associated with poorer n-back performance among participants in the reference sleep trajectory classes: persistently average longer sleep duration (B = −0.41, 95% CI −0.71 to −0.10, p = 0.009) and persistently average earlier MPS (B = −0.69, 95% CI −1.11 to −0.28, p = 0.001). No significant associations were observed between childhood sleep trajectory classes and working memory among participants without HS. Full results for all interaction models are reported in the Supplementary-Results.

**Table 4.** The interaction between sleep trajectories and experiencing hypomanic symptoms in association with N-back d′ at age 24^a^.

|  | **Model 1: Main effects** |  |  | **Model 2: Interaction effects** |  |  | **Model 3: Adjusted model** |  |  |
|  | **B** | **95% CI** | **p-value** | **B** | **95% CI** | **p-value** | **B** | **95% CI** | **p-value** |
|---|---|---|---|---|---|---|---|---|---|
| **Persistently Average Shorter Sleep Duration Class** | 0.05 | -0.14, 0.25 | 0.585 | 0.05 | -0.15, 0.25 | 0.617 | 0.08 | -0.11, 0.28 | 0.408 |
| Hypomanic Symptoms | -0.53 | -1.32, 0.26 | 0.192 | -0.64 | -1.66, 0.39 | 0.222 | -0.72 | -1.73, 0.29 | 0.160 |
| Persistently Average Shorter Sleep Duration Class x Hypomanic Symptoms | - | - | - | 0.27 | -1.34, 1.89 | 0.739 | 0.56 | -1.05, 2.17 | 0.493 |
| Child’s sex | - | - | - | - | - | - | -0.06 | -0.25, 0.14 | 0.581 |
| Child’s ethnicity | - | - | - | - | - | - | -0.66 | -1.14, -0.19 | 0.006 |
| Maternal Age at birth | - | - | - | - | - | - | 0.04 | 0.01, 0.06 | 0.001 |
| Gestational Age | - | - | - | - | - | - | 0.01 | -0.04, 0.06 | 0.680 |
| Family Adversity Index | - | - | - | - | - | - | 0.00 | -0.03, 0.03 | 0.909 |
| BPD traits at 11 years | - | - | - | - | - | - | -0.03 | -0.15, 0.10 | 0.684 |
| **Persistently Long Sleep Duration Class** | 0.01 | -0.12, 0.14 | 0.881 | -0.02 | -0.16, 0.11 | 0.750 | -0.07 | -0.20, 0.07 | 0.322 |
| Hypomanic Symptoms | -0.58 | -0.85, -0.32 | <0.001 | -0.74 | -1.03, -0.44 | <0.001 | -0.41 | -0.71, -0.10 | 0.009 |
| Persistently Long Sleep Duration Class x Hypomanic Symptoms | - | - | - | 0.83 | 0.15, 1.52 | 0.017 | 0.66 | -0.01, 1.34 | 0.053 |
| Child’s sex | - | - | - | - | - | - | -0.10 | -0.18, -0.01 | 0.022 |
| Child’s ethnicity | - | - | - | - | - | - | -0.33 | -0.56, -0.10 | 0.005 |
| Maternal Age at birth | - | - | - | - | - | - | 0.03 | 0.02, 0.04 | <0.001 |
| Gestational Age | - | - | - | - | - | - | 0.04 | 0.02, 0.06 | 0.001 |
| Family Adversity Index | - | - | - | - | - | - | -0.01 | -0.03, -0.00 | 0.016 |
| BPD traits at 11 years | - | - | - | - | - | - | -0.03 | -0.08, 0.01 | 0.166 |
| **Persistently Early MPS Class** | 0.01 | -0.08, 0.10 | 0.851 | 0.00 | -0.09, 0.09 | 0.996 | -0.02 | -0.11, 0.07 | 0.650 |
| Hypomanic Symptoms | -0.80 | -1.15, -0.46 | <0.001 | -0.98 | -1.38, -0.57 | <0.001 | -0.69 | -1.11, -0.28 | 0.001 |
|  | **Model 1: Main effects** |  |  | **Model 2: Interaction effects** |  |  | **Model 3: Adjusted model** |  |  |
|  | **B** | **95% CI** | **p-value** | **B** | **95% CI** | **p-value** | **B** | **95% CI** | **p-value** |
| Persistently Early MPS Class x Hypomanic Symptoms | - | - | - | 0.63 | -0.14, 1.40 | 0.111 | 0.71 | -0.05, 1.48 | 0.067 |
| Child’s sex | - | - | - | - | - | - | -0.11 | -0.19, -0.02 | 0.015 |
| Child’s ethnicity | - | - | - | - | - | - | -0.35 | -0.59, -0.10 | 0.006 |
| Maternal Age at birth | - | - | - | - | - | - | 0.03 | 0.02, 0.04 | <0.001 |
| Gestational Age | - | - | - | - | - | - | 0.03 | 0.01, 0.05 | 0.011 |
| Family Adversity Index | - | - | - | - | - | - | -0.01 | -0.02, 0.00 | 0.101 |
| BPD traits at 11 years | - | - | - | - | - | - | -0.04 | -0.09, 0.01 | 0.112 |
|  | **Model 1: Main effects** |  |  | **Model 2: Interaction effects** |  |  | **Model 3: Adjusted model** |  |  |
| **Term** | **B** | **95% CI** | **p-value** | **B** | **95% CI** | **p-value** | **B** | **95% CI** | **p-value** |
| **Persistently Average Shorter Sleep Duration Class** | 0.05 | -0.14, 0.25 | 0.585 | 0.05 | -0.15, 0.25 | 0.617 | 0.08 | -0.11, 0.28 | 0.408 |
| Hypomanic Symptoms | -0.53 | -1.32, 0.26 | 0.192 | -0.64 | -1.66, 0.39 | 0.222 | -0.72 | -1.73, 0.29 | 0.160 |
| Persistently Average Shorter Sleep Duration Class x Hypomanic Symptoms | - | - | - | 0.27 | -1.34, 1.89 | 0.739 | 0.56 | -1.05, 2.17 | 0.493 |
| Child’s sex | - | - | - | - | - | - | -0.06 | -0.25, 0.14 | 0.581 |
| Child’s ethnicity | - | - | - | - | - | - | -0.66 | -1.14, -0.19 | 0.006 |
| Maternal Age at birth | - | - | - | - | - | - | 0.04 | 0.01, 0.06 | 0.001 |
| Gestational Age | - | - | - | - | - | - | 0.01 | -0.04, 0.06 | 0.680 |
| Family Adversity Index | - | - | - | - | - | - | 0.00 | -0.03, 0.03 | 0.909 |
| BPD traits at 11 years | - | - | - | - | - | - | -0.03 | -0.15, 0.10 | 0.684 |
|  | **Model 1: Main effects** |  |  | **Model 2: Interaction effects** |  |  | **Model 3: Adjusted model** |  |  |
| **Term** | **B** | **95% CI** | **p-value** | **B** | **95% CI** | **p-value** | **B** | **95% CI** | **p-value** |
| **Persistently Long Sleep Duration Class** | 0.01 | -0.12, 0.14 | 0.881 | -0.02 | -0.16, 0.11 | 0.750 | -0.07 | -0.20, 0.07 | 0.322 |
| Hypomanic Symptoms | -0.58 | -0.85, -0.32 | <0.001 | -0.74 | -1.03, -0.44 | <0.001 | -0.41 | -0.71, -0.10 | 0.009 |
| Persistently Long Sleep Duration Class x Hypomanic Symptoms | - | - | - | 0.83 | 0.15, 1.52 | 0.017 | 0.66 | -0.01, 1.34 | 0.053 |
| Child’s sex | - | - | - | - | - | - | -0.10 | -0.18, -0.01 | 0.022 |
| Child’s ethnicity | - | - | - | - | - | - | -0.33 | -0.56, -0.10 | 0.005 |
| Maternal Age at birth | - | - | - | - | - | - | 0.03 | 0.02, 0.04 | <0.001 |
| Gestational Age | - | - | - | - | - | - | 0.04 | 0.02, 0.06 | 0.001 |
| Family Adversity Index | - | - | - | - | - | - | -0.01 | -0.03, -0.00 | 0.016 |
| BPD traits at 11 years | - | - | - | - | - | - | -0.03 | -0.08, 0.01 | 0.166 |
| **Persistently Early MPS Class** | 0.01 | -0.08, 0.10 | 0.851 | 0.00 | -0.09, 0.09 | 0.996 | -0.02 | -0.11, 0.07 | 0.650 |
| Hypomanic Symptoms | -0.80 | -1.15, -0.46 | <0.001 | -0.98 | -1.38, -0.57 | <0.001 | -0.69 | -1.11, -0.28 | 0.001 |
| Persistently Early MPS Class x Hypomanic Symptoms | - | - | - | 0.63 | -0.14, 1.40 | 0.111 | 0.71 | -0.05, 1.48 | 0.067 |
| Child’s sex | - | - | - | - | - | - | -0.11 | -0.19, -0.02 | 0.015 |
| Child’s ethnicity | - | - | - | - | - | - | -0.35 | -0.59, -0.10 | 0.006 |
| Maternal Age at birth | - | - | - | - | - | - | 0.03 | 0.02, 0.04 | <0.001 |
| Gestational Age | - | - | - | - | - | - | 0.03 | 0.01, 0.05 | 0.011 |
| Family Adversity Index | - | - | - | - | - | - | -0.01 | -0.02, 0.00 | 0.101 |
| BPD traits at 11 years | - | - | - | - | - | - | -0.04 | -0.09, 0.01 | 0.112 |

Note: B = unstandardised regression coefficient; CI = confidence interval; MPS=Midpoint of sleep
<sup>a</sup> All analyses were weighted for sex, ethnicity, maternal age, maternal socioeconomic status, preterm delivery, birthweight and family adversity.

### Sleep trajectories, hypomanic symptoms, and response inhibition

For response inhibition, persistently average shorter sleep duration was associated with poorer stop-signal performance (higher SSRT; B=11.19, 95% CI=7.08 to 15.30, p<0.001). The interaction between persistently average shorter sleep duration and HS was also significant. (B=36.55, 95% CI=8.33 to 64.78, p=0.011) (Table 5). In contrast, the interaction between persistently longer sleep duration and HS was associated with lower SSRTs (B=-125.21, 95% CI=-193.56 to −56.86, p<0.001). Persistently earlier MPS was not associated with SSRT, and no interaction with HS was observed. Full results for all interaction models are reported in the Supplementary-Results.

**Table 5.** The interaction between sleep trajectories and experiencing hypomanic symptoms in association with stop signal performance at age 24^a^.

|  | Model 1: Main effects |  |  | Model 2: Interaction effects |  |  | Model 3: Adjusted model |  |  |
|---|---|---|---|---|---|---|---|---|---|
|  | B | 95% CI | p-value | B | 95% CI | p-value | B | 95% CI | p-value |
|---|---|---|---|---|---|---|---|---|---|
| **Persistently Average Shorter Sleep Duration Class** | 16.43 | 12.35, 20.51 | <0.001 | 15.43 | 11.32, 19.55 | <0.001 | 11.19 | 7.08, 15.30 | <0.001 |
| Hypomanic Symptoms | 38.49 | 24.09, 52.88 | <0.001 | 17.51 | -1.41, 36.42 | 0.070 | 15.08 | -3.17, 33.33 | 0.105 |
| Persistently Average Shorter Sleep Duration Class x Hypomanic Symptoms | - | - | - | 49.65 | 20.55, 78.74 | <0.001 | 36.55 | 8.33, 64.78 | 0.011 |
| Child’s sex | - | - | - | - | - | - | 19.65 | 16.02, 23.28 | <0.001 |
| Child’s ethnicity | - | - | - | - | - | - | -12.43 | -22.38, -2.47 |  |
| Maternal Age at birth | - | - | - | - | - | - | 0.60 | 0.17, 1.02 | 0.014 |
| Gestational Age | - | - | - | - | - | - | 4.41 | 3.47, 5.35 | 0.006 |
| Family Adversity Index | - | - | - | - | - | - | 0.42 | -0.07, 0.91 | <0.001 |
| BPD traits at 11 years | - | - | - | - | - | - | 3.78 | 1.72, 5.84 | 0.095 |
| **Persistently Long Sleep Duration Class** | -3.36 | -11.59, 4.88 | 0.424 | -1.39 | -9.64, 6.86 | 0.741 | -1.93 | -10.03, 6.18 | 0.641 |
| Hypomanic Symptoms | 54.15 | 33.04, 75.26 | <0.001 | 68.32 | 46.16, 90.48 | <0.001 | 55.98 | 33.81, 78, 15 | <0.001 |
| Persistently Long Sleep Duration Class x Hypomanic Symptoms | - | - | - | -140.30 | -210.02, -70.58 | <0.001 | -125.21 | -193.56, -56.86 | <0.001 |
| Child’s sex | - | - | - | - | - | - | 16.82 | 11.75, 21.88 | <0.001 |
| Child’s ethnicity | - | - | - | - | - | - | -15.20 | -30.06, -0.34 | 0.045 |
| Maternal Age at birth | - | - | - | - | - | - | 0.40 | -0.18, 0.99 | 0.176 |
| Gestational Age | - | - | - | - | - | - | 3.37 | 2.06, 4.69 | <0.001 |
| Family Adversity Index | - | - | - | - | - | - | 0.52 | -0.15, 1.20 | 0.130 |
| BPD traits at 11 years | - | - | - | - | - | - | 4.31 | 1.36, 7.26 | 0.004 |
| **Persistently Early MPS Class** | 2.40 | -2.86, 7.67 | 0.371 | 2.57 | -2.72, 7.86 | 0.340 | 3.96 | -1.25, 9.17 | 0.136 |
|  | Model 1: Main effects |  |  | Model 2: Interaction effects |  |  | Model 3: Adjusted model |  |  |
|  | B | 95% CI | p-value | B | 95% CI | p-value | B | 95% CI | p-value |
| Hypomanic Symptoms | 54.73 | 33.59, 75.88 | <0.001 | 57.99 | 34.73, 81.26 | <0.001 | 47.71 | 24.70, 70.71 | <0.001 |
| Persistently Early MPS Class x Hypomanic Symptoms | - | - | - | -18.75 | -74.55, 37.06 | 0.510 | -26.27 | -80.73, 28.20 | 0.344 |
| Child’s sex | - | - | - | - | - | - | 17.76 | 12.67, 22.84 | <0.001 |
| Child’s ethnicity | - | - | - | - | - | - | -14.83 | -29.74, 0.08 | 0.051 |
| Maternal Age at birth | - | - | - | - | - | - | 0.32 | -0.27, 0.90 | 0.288 |
| Gestational Age | - | - | - | - | - | - | 3.17 | 1.84, 4.50 | <0.001 |
| Family Adversity Index | - | - | - | - | - | - | 0.61 | -0.07, 1.28 | 0.081 |
| BPD traits at 11 years | - | - | - | - | - | - | 4.72 | 1.75, 7.69 | 0.002 |

Note: B = unstandardised regression coefficient; CI = confidence interval; MPS=Midpoint of sleep
<sup>a</sup> All analyses were weighted for sex, ethnicity, maternal age, maternal socioeconomic status, preterm delivery, birthweight and family adversity.

## Discussion

Using longitudinal data from ALSPAC, a large UK population-based cohort, we identified distinct developmental trajectories of night-time sleep duration and MPS from infancy to late childhood. We found that persistently average shorter sleep duration and persistently longer sleep duration during childhood were each associated with an increased likelihood of experiencing HS at ages 21–23, whereas persistently earlier MPS was associated with a lower likelihood of experiencing HS. Interaction analyses showed that persistently average shorter sleep duration in childhood, in combination with HS at ages 21–23 years, was associated with poorer response inhibition at age 24, whereas persistently longer sleep duration in childhood, in combination with HS, was associated with better response inhibition.

First, we found that both persistently average shorter sleep duration and persistently longer sleep duration were associated with an increased risk of experiencing HS in early adulthood. The association between persistently shorter sleep duration and HS could not be reliably estimated, as only 16 participants in this class had available HS data, none of whom met the criteria for HS. Consequently, the findings for this class should be interpreted in light of the limited sample size and sparse outcome data. Notably, the association with persistently average shorter sleep duration with an increased risk of experiencing HS in early adulthood suggests that even relatively modest, sustained differences in childhood sleep duration may be relevant to experiencing later potential hypomanic symptoms. Persistently average shorter sleep duration may reflect heightened arousal, altered sleep need, or early-emerging difficulties in sleep–wake regulation, processes that have been implicated in later hypomanic and manic symptomatology [40]. In contrast, persistently longer sleep duration may reflect a distinct vulnerability phenotype characterised by altered energy regulation [41]. Evidence from clinical and high-risk studies indicates that hypersomnia and prolonged sleep are common features of bipolar-spectrum disorders [2], observed across mood states and BD subtypes [42]. These studies may reflect differing reasons for the associations that we found, but the present study cannot further establish their underlying mechanisms.

Second, we found that persistently earlier MPS in childhood was associated with a lower likelihood of experiencing HS, suggesting that earlier circadian timing may confer resilience to experiencing HS. Although chronotype reflects an inter-individual variation in circadian timing and is not inherently pathological [43], at-risk BD populations do demonstrate eveningness chronotype and social and circadian dysrhythmias [2]. From a neurobiological perspective, a persistently earlier MPS may reflect stronger circadian entrainment and reduced circadian misalignment across development, potentially facilitating more stable sleep–wake regulation [44,45]. Our findings raise the possibility that an earlier MPS during childhood may reduce susceptibility to experiencing HS by potentially supporting more robust circadian regulation, although this requires further study.

Third, we found that persistently average shorter sleep duration across childhood was associated with poorer response inhibition, a measure of inhibitory control, at 24 years. Moreover, the combination of persistently average shorter sleep duration and HS was associated with poorer inhibitory control than either risk factor alone. One possible explanation is that altered frontostriatal functioning, which is implicated in response inhibition, may contribute to the observed association between childhood sleep trajectories, hypomanic symptoms, and inhibitory control [46,47]. Previous research has linked altered frontostriatal functional connectivity to impaired response inhibition in individuals with insomnia disorder [47], while frontostriatal dysfunction has also been implicated in impulsivity associated with BD [46,48]. Taken together, these findings raise the possibility that persistently shorter sleep duration across childhood may confer vulnerability in inhibitory control systems that becomes more apparent when HS are also present. However, these interpretations remain speculative given the modest effect sizes we found and this requires further examination. In contrast, individuals with both persistently longer sleep duration in childhood and HS demonstrated comparatively better inhibitory performance. This unexpected finding should be interpreted cautiously. Replication in independent cohorts with objective sleep measures, repeated assessments of mood symptoms, and neurocognitive follow-up is needed to assess its reproducibility. Although we found that persistently earlier MPS was associated with lower odds of HS, we did not find an association with response inhibition at age 24. However, as sleep timing and circadian phase were not assessed concurrently with cognitive performance, we could not determine whether current circadian characteristics were associated with differences in inhibitory control [49].

Fourth, in contrast to response inhibition findings, the childhood sleep trajectories that we found to be associated with HS were not associated with working memory performance at age 24, nor did they moderate the association between HS and working memory. However, HS reported at ages 21–23 were associated with poorer n-back performance at age 24 among participants in the persistently average longer sleep-duration class and the persistently average earlier MPS class. As HS were not assessed concurrently with cognitive performance, we could not determine whether these associations reflected ongoing mood symptoms or more enduring differences in working memory.

### Strengths and Limitations

Strengths of our study include the large population-based sample size and a developmental approach integrating longitudinal methodology. Another strength is that the exposure (sleep at 6 months to 11 years) and outcomes (at age 21-23 and 24 years) were temporally separated, which reduces the likelihood of reverse causality of the outcome.

However, there are also several limitations. First, HS were assessed at a single time point at ages 21–23 using the self-reported HCL-32, which asks participants to recall lifetime symptoms. Without repeated assessments or clinical verification, we could not examine the stability or reliability of HS measured by the HCL-32 over time, and recall or classification errors are quite possible. Further, as the age of onset of HS was unavailable, we could not establish whether these symptoms first emerged after the childhood sleep assessments were completed. Second, sleep measures were based on parental report rather than objective assessments such as actigraphy, potentially introducing reporting bias. The trajectories were also based on night-time sleep duration only, without accounting for daytime napping. Our analyses also did not capture other potentially relevant sleep and circadian features, including sleep quality, rhythm stability, variability, circadian amplitude or social rhythmicity, as these were not assessed at the relevant ALSPAC time points. Our trajectories were also limited to childhood, yet circadian phase shifts substantially across adolescence and young adulthood. Cognitive performance at age 24 may also have been influenced by contemporaneous sleep disturbances, which were not accounted for in our analyses. Third, as with any observational analysis, residual confounding remains possible. Although we adjusted for several demographic and early-life factors selected a priori, other potentially relevant characteristics, including parental history of BD, childhood attention deficit/hyperactivity disorder (ADHD) symptoms, body mass index (BMI), chronic physical illness and health-related behaviours, were not included in our models. Fourth, although inverse probability weighting partially addressed cohort-specific patterns of non-response, bias from unmeasured factors that could additionally influence missingness cannot be dismissed. Finally, the ALSPAC cohort comprises mainly White participants, reducing the generalisability of our findings to other ethnicities.

### Conclusions

Findings from this study suggest that both persistently average shorter and persistently longer sleep duration across childhood are associated with increased risk of HS in young adulthood, whereas persistently earlier MPS is associated with a lower likelihood. Our findings also suggest that shorter sleep duration in childhood may be associated with poorer response inhibition among individuals who have experienced HS, whereas persistently longer sleep duration may be associated with better response inhibition. Together, these results underscore the potential importance of developmental sleep and circadian regulation in understanding the development, and potential prevention, of hypomanic and cognitive dysfunction symptoms in young adults.

## Supporting information

Supplementary Methods, Results, Tables and Figures

## Data Availability

Data was obtained from the ALSPAC. Please note that the study website contains details of all the data that is available through a fully searchable data dictionary and variable search tool: http://www.bristol.ac.uk/alspac/researchers/our-data/. Any research team can apply to use the ALSPAC and submit a protocol with an accompanying research question. It is not within the rights of the authors to share this data as it requires individual ALSPAC approvals per researcher.

## Acknowledgements

We are extremely grateful to all the families who took part in this study, the midwives for their help in recruiting them, and the whole ALSPAC team, which includes data collection staff, data and administrations staff, technical managers and the technical staff with the Bristol Bioresource Laboratory based within the University of Bristol.

## Authors contributions

Durdurak, B.B. - Formal analysis, data validation, writing – original draft, and writing – review & editing, methodology, visualisation, project administration. Marwaha, S. – Conceptualization, funding acquisition, supervision, resources, software, writing – review & editing. Stokes, PRA - Conceptualisation, formal analyses, supervision, validation, methodology, writing – review & editing. Morales-Muñoz, I. - Conceptualisation, data curation, formal analyses, funding acquisition, supervision, validation, methodology, resources, software, project administration, writing – review & editing. The corresponding author had final responsibility for the decision to submit for publication.

## References

[1] Durdurak BB, Altaweel N, Upthegrove R, Marwaha S. Understanding the development of bipolar disorder and borderline personality disorder in young people: a meta-review of systematic reviews. Psychol Med 2022;52:3769–82. 10.1017/S0033291722003002.

[2] Scott J, Etain B, Miklowitz D, Crouse JJ, Carpenter J, Marwaha S, et al. A systematic review and meta-analysis of sleep and circadian rhythms disturbances in individuals at high-risk of developing or with early onset of bipolar disorders. Neurosci Biobehav Rev 2022;135:104585. 10.1016/j.neubiorev.2022.104585.

[3] De Crescenzo F, Economou A, Sharpley AL, Gormez A, Quested DJ. Actigraphic features of bipolar disorder: A systematic review and meta-analysis. Sleep Med Rev 2017;33:58–69. 10.1016/j.smrv.2016.05.003.

[4] Scott J, Murray G, Henry C, Morken G, Scott E, Angst J, et al. Activation in bipolar disorders a systematic review. JAMA Psychiatry 2017;74:189–96. 10.1001/jamapsychiatry.2016.3459.

[5] Melo MCA, Garcia RF, Linhares Neto VB, Sá MB, de Mesquita LMF, de Araújo CFC, et al. Sleep and circadian alterations in people at risk for bipolar disorder: A systematic review. J Psychiatr Res 2016;83:211–9. 10.1016/j.jpsychires.2016.09.005.

[6] Van Meter AR, Burke C, Youngstrom EA, Faedda GL, Correll CU. The Bipolar Prodrome: Meta-Analysis of Symptom Prevalence Prior to Initial or Recurrent Mood Episodes. J Am Acad Child Adolesc Psychiatry 2016;55:543–55. 10.1016/j.jaac.2016.04.017.

[7] Ritter PS, Höfler M, Wittchen HU, Lieb R, Bauer M, Pfennig A, et al. Disturbed sleep as risk factor for the subsequent onset of bipolar disorder - Data from a 10-year prospective-longitudinal study among adolescents and young adults. J Psychiatr Res 2015;68:76–82. 10.1016/j.jpsychires.2015.06.005.

[8] Dondé C, Jaffiol A, Khouri C, Pouchon A, Tamisier R, Lejoyeux M, et al. Sleep disturbances in early clinical stages of psychotic and bipolar disorders: A meta-analysis. Australian and New Zealand Journal of Psychiatry 2022;56:1068–79. 10.1177/00048674211068395.

[9] Pancheri C, Verdolini N, Pacchiarotti I, Samalin L, Chiaie RD, Biondi M. A systematic review on sleep alterations anticipating the onset of bipolar disorder. European Psychiatry 2019;58:45–53. 10.1016/j.eurpsy.2019.02.003.

[10] Morales-Muñoz I, Marwaha S, Upthegrove R, Cropley V. Role of Inflammation in Short Sleep Duration Across Childhood and Psychosis in Young Adulthood. JAMA Psychiatry 2024:1–9. 10.1001/jamapsychiatry.2024.0796.

[11] Hosang GM, Martin J, Karlsson R, Lundström S, Larsson H, Ronald A, et al. Association of Etiological Factors for Hypomanic Symptoms, Bipolar Disorder, and Other Severe Mental Illnesses. JAMA Psychiatry 2022;79:143–50. 10.1001/jamapsychiatry.2021.3654.

[12] Grande I, Berk M, Birmaher B, Vieta E. Bipolar disorder. The Lancet 2016;387:1561–72. 10.1016/S0140-6736(15)00241-X.

[13] Nieuwenhuys A, Wadsley CG, Sullivan R, Cirillo J, Byblow WD. Tired and out of control? Effects of total and partial sleep deprivation on response inhibition under threat and no-threat conditions. Sleep 2025;48:1–13. 10.1093/sleep/zsae275.

[14] Pasula EY, Brown GG, McKenna BS, Mellor A, Turner T, Anderson C, et al. Effects of sleep deprivation on component processes of working memory in younger and older adults. Sleep 2018;41:1–9. 10.1093/sleep/zsx213.

[15] Zhao R, Zhang X, Fei N, Zhu Y, Sun J, Liu P, et al. Decreased cortical and subcortical response to inhibition control after sleep deprivation. Brain Imaging Behav 2019;13:638–50. 10.1007/s11682-018-9868-2.

[16] Könen T, Dirk J, Schmiedek F. Cognitive benefits of last night’s sleep: Daily variations in children’s sleep behavior are related to working memory fluctuations. J Child Psychol Psychiatry 2015;56:171–82. 10.1111/jcpp.12296.

[17] Xie W, Berry A, Lustig C, Deldin P, Zhang W. Poor Sleep Quality and Compromised Visual Working Memory Capacity. Journal of the International Neuropsychological Society 2019;25:583–94. 10.1017/S1355617719000183.

[18] Pearson O, Uglik-Marucha N, Miskowiak KW, Cairney SA, Rosenzweig I, Young AH, et al. The relationship between sleep disturbance and cognitive impairment in mood disorders: A systematic review. J Affect Disord 2023;327:207–16. 10.1016/j.jad.2023.01.114.

[19] Sankar A, Ziersen SC, Ozenne B, Beaman EE, Dam VH, Fisher PM, et al. Association of neurocognitive function with psychiatric hospitalization and socio-demographic conditions in individuals with bipolar and major depressive disorders. EClinicalMedicine 2023;58:101927. 10.1016/j.eclinm.2023.101927.

[20] Van Rheenen TE, Lewandowski KE, Bauer IE, Kapczinski F, Miskowiak K, Burdick KE, et al. Current understandings of the trajectory and emerging correlates of cognitive impairment in bipolar disorder: An overview of evidence. Bipolar Disord 2020;22:13–27. 10.1111/bdi.12821.

[21] Miskowiak KW, Kjærstad HL, Meluken I, Petersen JZ, Maciel BR, Köhler CA, et al. The search for neuroimaging and cognitive endophenotypes: A critical systematic review of studies involving unaffected first-degree relatives of individuals with bipolar disorder. Neurosci Biobehav Rev 2017;73:1–22. 10.1016/j.neubiorev.2016.12.011.

[22] Burdick KE, Millett CE. Cognitive heterogeneity is a key predictor of differential functional outcome in patients with bipolar disorder. European Neuropsychopharmacology 2021;53:4–6. 10.1016/j.euroneuro.2021.06.008.

[23] Burgess JL, Bradley AJ, Anderson KN, Gallagher P, McAllister-Williams RH. The relationship between physical activity, BMI, circadian rhythm, and sleep with cognition in bipolar disorder. Psychol Med 2022;52:467–75. 10.1017/S003329172000210X.

[24] Bradley AJ, Anderson KN, Gallagher P, McAllister-Williams RH. The association between sleep and cognitive abnormalities in bipolar disorder. Psychol Med 2020;50:125–32. 10.1017/S0033291718004038.

[25] Laskemoen JF, Büchmann C, Barrett EA, Collier-Høegh M, Haatveit B, Vedal TJ, et al. Do sleep disturbances contribute to cognitive impairments in schizophrenia spectrum and bipolar disorders? Eur Arch Psychiatry Clin Neurosci 2020;270:749–59. 10.1007/s00406-019-01075-0.

[26] Boyd A, Golding J, Macleod J, Lawlor DA, Fraser A, Henderson J, et al. Cohort profile: The ‘Children of the 90s’-The index offspring of the avon longitudinal study of parents and children. Int J Epidemiol 2013;42:111–27. 10.1093/ije/dys064.

[27] Fraser A, Macdonald-wallis C, Tilling K, Boyd A, Golding J, Davey smith G, et al. Cohort profile: The avon longitudinal study of parents and children: ALSPAC mothers cohort. Int J Epidemiol 2013;42:97–110. 10.1093/ije/dys066.

[28] Northstone K, Ben Shlomo Y, Teyhan A, Hill A, Groom A, Mumme M, et al. The Avon Longitudinal Study of Parents and children ALSPAC G0 Partners: A cohort profile. Wellcome Open Res 2023;8:37. 10.12688/wellcomeopenres.18782.2.

[29] Simpkin CT, Jenni OG, Carskadon MA, Wright KP, Akacem LD, Garlo KG, et al. Chronotype is associated with the timing of the circadian clock and sleep in toddlers. J Sleep Res 2014;23:397–405. 10.1111/jsr.12142.

[30] Kantermann T, Sung H, Burgess HJ. Comparing the Morningness-Eveningness Questionnaire and Munich ChronoType Questionnaire to the dim light melatonin onset. J Biol Rhythms 2015;30:449–53. 10.1177/0748730415597520.

[31] Angst J, Cassano G. The mood spectrum: Improving the diagnosis of bipolar disoder. Bipolar Disorders, Supplement 2005;7:4–12. 10.1111/j.1399-5618.2005.00210.x.

[32] Kirchner WK. AGE DIFFERENCES IN SHORT-TERM RETENTION OF RAPIDLY CHANGING INFORMATION Minnesota Mining and Manufacturing Company. J Exp Psychol 1958;55:352–8.

[33] Logan GD, Cowan WB, Davis KA. On the ability to inhibit simple and choice reaction time responses: A model and a method. J Exp Psychol Hum Percept Perform 1984;10:276–91. 10.1037/0096-1523.10.2.276.

[34] Zanarini MC. Zanarini Rating Scale for Borderline Personality Disorder (ZAN-BPD): A continuous measure of DSM-IV borderline psychopathology. J Pers Disord 2003;17:233–42. 10.1521/pedi.17.3.233.22147.

[35] Morales-Muñoz I, Broome MR, Marwaha S. Association of Parent-Reported Sleep Problems in Early Childhood with Psychotic and Borderline Personality Disorder Symptoms in Adolescence. JAMA Psychiatry 2020;77:1256–65. 10.1001/jamapsychiatry.2020.1875.

[36] Winsper C, Wolke D, Scott J, Sharp C, Thompson A, Marwaha S. Psychopathological outcomes of adolescent borderline personality disorder symptoms. Australian and New Zealand Journal of Psychiatry 2020;54:308–17. 10.1177/0004867419882494.

[37] Muthén B, Muthén L. Mplus: A General Latent Variable Modeling Program 2017.

[38] Jung T, Wickrama KAS. An Introduction to Latent Class Growth Analysis and Growth Mixture Modeling. Soc Personal Psychol Compass 2008;2:302–17. 10.1111/j.1751-9004.2007.00054.x.

[39] Wardenaar KJ. Latent class growth analysis and growth mixture modeling using R: A tutorial for two R-packages and a comparison with Mplus. PsyArXiv 2022:1–23.

[40] Hensch T, Wozniak D, Spada J, Sander C, Ulke C, Wittekind DA, et al. Vulnerability to bipolar disorder is linked to sleep and sleepiness. Transl Psychiatry 2019;9:1–10. 10.1038/s41398-019-0632-1.

[41] Mansur RB, Lee Y, McIntyre RS, Brietzke E. What is bipolar disorder? A disease model of dysregulated energy expenditure. Neurosci Biobehav Rev 2020;113:529–45. 10.1016/j.neubiorev.2020.04.006.

[42] Grigolon RB, Trevizol AP, Cerqueira RO, Lee Y, Mansur RB, McIntyre RS, et al. Hypersomnia and Bipolar Disorder: A systematic review and meta-analysis of proportion. J Affect Disord 2019;246:659–66. 10.1016/j.jad.2018.12.030.

[43] Walker M. Why we sleep: Unlocking the power of sleep and dreams. Penguin Books Ltd; 2017.

[44] Li SX, Chan NY, Man Yu MW, Lam SP, Zhang J, Yan Chan JW, et al. Eveningness chronotype, insomnia symptoms, and emotional and behavioural problems in adolescents. Sleep Med 2018;47:93–9. 10.1016/j.sleep.2018.03.025.

[45] Mondin TC, Cardoso T de A, Souza LD de M, Jansen K, da Silva Magalhães PV, Kapczinski F, et al. Mood disorders and biological rhythms in young adults: A large population-based study. J Psychiatr Res 2017;84:98–104. 10.1016/j.jpsychires.2016.09.030.

[46] Chan CC, Alter S, Hazlett EA, Shafritz KM, Yehuda R, Goodman M, et al. Neural correlates of impulsivity in bipolar disorder: A systematic review and clinical implications. Neurosci Biobehav Rev 2023;147:105109. 10.1016/j.neubiorev.2023.105109.

[47] Zhang H, Sun H, Li J, Yang J, Fan Y, Jülich ST, et al. Response inhibition impairment related to altered frontal-striatal functional connectivity in insomnia disorder: A pilot and non-clinical study. J Psychiatr Res 2024;170:138–46. 10.1016/j.jpsychires.2023.12.023.

[48] Hummer TA, Hulvershorn LA, Karne HS, Gunn AD, Wang Y, Anand A. Emotional response inhibition in bipolar disorder: A functional magnetic resonance imaging study of trait- and state-related abnormalities. Biol Psychiatry 2013;73:136–43. 10.1016/j.biopsych.2012.06.036.

[49] Song J, Feng P, Zhao X, Xu W, Xiao L, Zhou J, et al. Chronotype regulates the neural basis of response inhibition during the daytime. Chronobiol Int 2018;35:208–18. 10.1080/07420528.2017.1392550.

