## Supplementary Methods, Results, Tables and Figures for "Childhood sleep trajectories, hypomanic symptoms, and cognitive functioning in early adulthood: a longitudinal cohort study"

**S1. Study population**

**S2. Measures**
S2.1 Midpoint of sleep
S2.2 Hypomanic symptoms
S2.3 Cognitive outcomes: working memory and response inhibition
S2.4 Confounders

**S3. Statistical analyses**
S3.1 Latent class growth analysis and model selection
S3.2 Selection of the night-time sleep duration class solution
S3.3 Selection of the midpoint of sleep class solution
S3.4 Inverse probability weighting
S3.5 Cross-tabulations for sparse cells/separation

**Supplementary Results**

**S4. Additional information on interaction models**
S4.1 Working memory models
S4.2 Response inhibition models

**Supplementary Tables and Figures**

**Table S1.** Cross-tabulation of night-time sleep duration trajectory classes and clinically significant hypomanic symptoms.

**Table S2.** Cross-tabulation of MPS trajectory classes and clinically significant hypomanic symptoms.

**Table S3.** Differences in socio-demographic variables between non-participating and participating subjects in the study for hypomanic symptoms at age 21-23 years

**Table S4**. Differences in socio-demographic variables between non-participating and participating subjects in the study for the Stop Signal test at age 24 years

**Table S5.** Differences in socio-demographic variables between non-participating and participating subjects in the study for N-Back test at age 24 years

**Figure S1.** Attrition of participants from the ALSPAC cohort

**Supplementary References**

### S1. Study population

The Avon Longitudinal Study of Parents and Children (ALSPAC) is a UK birth cohort study examining the determinants of development, health, and disease during childhood and beyond. Pregnant women resident in Avon, UK with expected dates of delivery between 1st April 1991 and 31st December 1992 were invited to take part in the study. 20,248 pregnancies have been identified as being eligible and the initial number of pregnancies enrolled was 14,541 (Boyd et al., 2013; Fraser et al., 2013). Of the initial pregnancies, there was a total of 14,676 foetuses, resulting in 14,062 live births and 13,988 children who were alive at 1 year of age. When the oldest children were approximately 7 years of age, an attempt was made to bolster the initial sample with eligible cases who had failed to join the study originally. As a result, when considering variables collected from the age of seven onwards (and potentially abstracted from obstetric notes) there are data available for more than the 14,541 pregnancies mentioned above: The number of new pregnancies not in the initial sample (known as Phase I enrolment) that are currently represented in the released data and reflecting enrolment status at the age of 24 is 906, resulting in an additional 913 children being enrolled (456, 262 and 195 recruited during Phases II, III and IV respectively). The phases of enrolment are described in more detail in the cohort profile paper and its update (see footnote 5 below). The total sample size for analyses using any data collected after the age of seven is therefore 15,447 pregnancies, resulting in 15,658 foetuses. Of these 14,901 children were alive at 1 year of age. From the age of 30, there were an additional 101 new participants recruited from 101 pregnancies during Phase V enrolment. The total sample size for analyses using any data collected after the age of 30 is therefore 15,548 pregnancies, resulting in 15,690 foetuses. Of these 15,002 children were alive at 1 year of age.

Of the original 14,541 initial pregnancies, 338 were from a woman who had already enrolled with a previous pregnancy, meaning 14,203 unique mothers were initially enrolled in the study. As a result of the additional phases of recruitment, a further 630 women who did not enrol originally have provided data since their child was 7 years of age. This provides a total of 14,833 unique women (G0 mothers) enrolled in ALSPAC as of September 2021.

G0 partners were invited to complete questionnaires by the mothers at the start of the study and they were not formally enrolled at that time. 12,113 G0 partners have been in contact with the study by providing data and/or formally enrolling when this first started in 2010. 3,807 G0 partners are currently enrolled.

Study data were collected and managed using REDCap electronic data capture tools hosted at the University of Bristol (Harris et al., 2009). REDCap (Research Electronic Data Capture) is a secure, web-based software platform designed to support data capture for research studies.

Ethical approval for the study was obtained from the ALSPAC Ethics and Law Committee and the Local Research Ethics Committees. Informed consent for the use of all data collected was obtained from participants following the recommendations of the ALSPAC Ethics and Law Committee at the time. Participants can contact the study team at any time to retrospectively withdraw consent for their data to be used. Study participation is voluntary and during all data collection sweeps, information was provided on the intended use of data.

The completion of a questionnaire, either on paper or online, was considered to be written consent from participants to use their data for research purposes.

For the majority of tests undertaken during face to face visits, verbal consent was obtained from participants (both parents and children as appropriate) prior to the start of any data collection. However, some tests required the completion of a written consent form.

Please note that the study website contains details of all the data that is available through a fully searchable data dictionary and variable search tool" and reference the following webpage: http://www.bristol.ac.uk/alspac/researchers/our-data/.

### S2. Measures

#### *S2.1 Midpoint of sleep*

MPS shows a strong association with dim-light melatonin onset in both toddlers (Simpkin et al., 2014) and adults (Kantermann et al., 2015). Moreover, MPS has been shown to be a more robust indicator of chronotype than bedtime alone across age groups, including adults (Kantermann et al., 2015), children (Werner et al., 2009), and toddlers (Simpkin et al., 2014). On this basis, both bedtime and MPS were included in the present study as indirect markers of chronotype. The use of MPS from as early as 6 months of age is supported by prior evidence indicating that circadian regulation of cortisol emerges between 2 weeks and 9 months in infancy (Ivars et al., 2015), and that by approximately 2 months of age, clear circadian patterns are observable in both sleep–wake behaviour and hormonal secretion (Rivkees, 2003).

#### *S2.2 Hypomanic symptoms*

The HCL-32 is a self-reported questionnaire that assesses lifetime history of hypomanic symptoms. It includes detailed assessments of bipolar mood, energy, and activity levels (in total 32 items). The instrument has been used extensively and validated in several studies (Angst et al., 2011; Forty et al., 2009; Meyer et al., 2007, 2014a). In clinical settings, it is considered to be a clinically useful screening instrument for hypomania and bipolar disorder type II.(Smith et al., 2015) In ALSPAC, when the cohort were 21-23 years of age, postal and online questionnaires for assessing lifetime history of hypomanic symptoms, using HCL-32 were sent. The HCL-32 has been used as both a continuous and categorical measure of hypomanic symptoms (Carvalho et al., 2015; Court et al., 2014) Respondents were asked to consider a time when they were in a “high or hyper” condition and endorse statements about their emotions, thoughts, and behaviour during this time. Although initially the HCL-32 was developed as a screening instrument for people with BD type II in people with recurrent depressive disorders, it is also a valid and sensitive tool for young, nonclinical populations (Meyer et al., 2014b). In our study, we used a categorical measure. Hypomanic symptoms were not clinically verified in the cohort.

*S2.3 Cognitive outcomes: working memory and response inhibition*

**Working memory:** An n-back task was used as a continuous performance measure of working memory (Kirchner, 1958). Participants monitored a series of numbers and indicated whether each number matched the one they saw two trials previously. Participants responded via keypress. The task consisted of 48 trials, eight of which were target trials (i.e. matches). The primary outcome measure was d prime (d’), a discriminability index that takes into account the proportion of hits (correctly identified matches) to false alarms (non-matches incorrectly identified as matches) to estimate signal-detection ability (McNichol, 1972). Higher d’ indicates better performance. Participants were excluded if they responded to fewer than 50% of trials or had a negative d’.

**Response inhibition:** A stop signal task was used to assess participants' capacity to withhold a motor response (Logan et al., 1984). Participants were presented with a series of trials displaying either an ‘X’ or ‘O’ on a blank screen. On each trial, participants were asked to respond by pressing the corresponding key (X or O) as quickly as possible, unless they heard an auditory tone (the ‘stop signal’). Participants completed four blocks of 64 trials, 25% of which had a stop signal. The delay between stimulus onset and stop signal (stop signal delay) varied between trials. The primary outcome was stop signal reaction time (SSRT), calculated as the difference between the median reaction time for go trials and an estimate of the median stop signal delay (SSRT = Go Reaction Timemed – Median Stop Signal Delay). Median Stop Signal Delay was the latency where each participant was likely to fail to inhibit 50% of trials. Lower SSRTs indicate better response inhibition.

*S2.4 Confounders*

Covariates were defined a priori based on developmental and epidemiological evidence indicating that early-life exposures shape both sleep regulation (Kajeepeta et al., 2015) and vulnerability to BD (Palmier-Claus et al., 2016). Specifically, covariates included child’s sex at birth, given sex differences in BD subtype prevalence (Dell’Osso et al., 2021); child’s ethnicity, due to variation in BD prevalence across ethnic groups (Clemente et al., 2015); mother’s age at birth, given associations between advanced maternal age and later BD risk (Polga et al., 2022); gestational age, given that early gestational age has been linked to later BD (Nosarti et al., 2012); and early family adversity, including socioeconomic disadvantage, maternal psychopathology, and adverse family conditions, given associations with later BD risk (Bergink et al., 2016; Palmier-Claus et al., 2016). Borderline personality disorder (BPD) traits at age 11 were additionally included, as these traits are strongly associated with both sleep disturbances and hypomanic symptoms (Morales-Muñoz et al., 2020; Winsper et al., 2020).

Child’s sex, ethnicity, mother’s age at birth and gestational age were reported by the mother. Early-life adversity including but not limited to family psychopathology, socioeconomic status and childhood abuse were assessed using the Family Adversity Index (FAI) during pregnancy and at 2 and 4 years. BPD traits at age 11 were assessed via the Childhood Interview for DSM-IV Borderline Personality Disorder: UK Version (CI-BPD-UK), a face-to-face semi-structured interview based on the borderline module of the Diagnostic Interview for DSM-IV Personality Disorders (Zanarini, 2003).

***Adverse Childhood Experiences using Family Adversity Index*:**

Family Adversity Index (FAI) total score was obtained using FAI index during pregnancy, and at 2 and 4 years, which includes items on (a)death or illness in family, (b) child's experience of violent victimization (e.g., physical or sexual abuse), (c) inter-parental conflict, (d) family disruption (e.g., mother got divorced or separated), (e) parental employment difficulties, (f) parental legal difficulties, (g) parental psychopathology (e.g., mother or father reported high levels of depression or anxiety symptomatology), (h) parental substance use(e.g., mother or father reported heavy alcohol consumption or illegal drug use), (i) financial hardship, and (j) housing inadequacies or instability.

***Borderline Personality Disorder Symptoms:***

Borderline Personality Disorder (BPD) was evaluated through a face-to-face semi-structured interview: the UK Childhood Interview for DSM-IV Borderline Personality Disorder (UK-CI-BPD; Zanarini, 2003). This assessment tool is derived from the borderline module of the Diagnostic Interview for DSM-IV Personality Disorders (DIPD-IV; Zanarini, 2003), a widely utilized semi-structured interview designed for assessing all DSM-IV Axis II disorders. The inter-rater and test–retest reliability of the DSM-III, DSM-III-R and DSM-IV versions of this measure have all proven to be good to excellent (Zanarini & Frankenburg, 2001). It is the first semi-structured interview assessing DSM-IV BPD in children and adolescents (Winsper, Hall, et al., 2017; Winsper, Tang, et al., 2017). The interview covers nine sections: intense inappropriate anger, affective instability, emptiness, identity disturbance, paranoid ideation/dissociation, frantic efforts to avoid abandonment, suicidal or self-mutilating behaviours, general impulsivity, and intense unstable relationships (Wolke et al., 2012). If the symptom occurred daily or approximately 25% of the time, the symptom was classed as definitely present; and probable if it had occurred repeatedly but did not meet criterion for definitely present.

### S3. Statistical analyses

#### *S3.1 Latent class growth analysis and model selection*

Model fit was assessed using the following parameters sample size–adjusted Bayesian information criterion, Lo-Mendell- Rubin likelihood ratio test, Vuong-Lo-Mendell-Rubin test, entropy value and proportion of individuals in each class. In line with previous studies utilising LCGA methodology, ABIC was given priority instead of Bayesian Information Criteria (BIC) because of previous studies indicating that it outperforms other information criteria indices such as Akaike Information Criteria (AIC) and Bayesian Information Criteria (BIC; Jones et al., 2018; Morgan, 2015; Paksarian et al., 2016; Tofighi & Enders, 2008), especially when small classes are present. Lower BIC values suggest better model fit. A significant VLMR value suggests that a k-class model fits the data better than a $K - 1$ class model. Entropy, a measure of the quality of classification, was additionally used to select the best model fit; an entropy value close to 1 is ideal. Finally, to decide the optimal class solution, an emphasis was also placed on proportion of individuals in each class, distinctiveness, and clinical relevance of the classes.

*S3.2 Selection of the night-time sleep duration class solution*

Determining the number of night-time sleep duration classes. Model selection was based on a combination of statistical fit, classification quality, class size, replication, and clinical interpretability, rather than on any single index. The Bayesian information criterion decreased with each added class, a pattern expected in large samples and one that does not, by itself, identify an optimal solution. The VLMR and LMR-adjusted likelihood ratio tests were statistically significant for the two-, three-, and four-class models, indicating that each successive solution up to four classes improved on the preceding one.

Although the likelihood ratio tests were nominally significant for the five-class model, this solution was not retained because it was not statistically admissible. The best log-likelihood was not replicated across random starts, indicating convergence on a local rather than global maximum, and the parametric bootstrapped likelihood ratio test was correspondingly flagged as unreliable. The fifth class comprised approximately seven participants (<0.1% of the sample) with an estimated mean night-time sleep duration of around 32 hours, a physiologically implausible value reflecting a small number of extreme outliers rather than a substantive developmental trajectory. The marginally higher entropy of the five-class model was an artefact of the near-perfect separation of these outliers and does not indicate superior model fit. The four-class solution, by contrast, converged to a replicated global maximum, contained no degenerate or implausible classes, and yielded four clinically interpretable trajectories, each comprising a sufficient proportion of the sample (≥3.1%).

The four-class solution was preferred over the three-class solution on substantive as well as statistical grounds. The three-class model collapsed the central region of the distribution into a single broad "average" trajectory, whereas the four-class model distinguished a persistently average-shorter trajectory from the average-longer majority. The four-class model therefore captured a developmentally and clinically meaningful gradient in sleep duration that the three-class model did not.

*S3.3 Selection of the midpoint of sleep class solution*

VLMR and LMRALT showed a statistically significant difference for the 2-class, 3-class, and 4-class models. Although the 5- and 6-class models had higher entropy values, they did not offer a significantly better fit than the 3- and 4-class models, respectively. Finally, the 3-class model reported a higher entropy value (0.648) than the 4-class model (0.643). Further, all classes from the 3-class model included sufficient sample sizes and were clinically relevant.

*S3.4 Inverse probability weighting*

It is possible that selection may have induced bias due to non- random missingness of data in ALSPAC (Weavers et al., 2021) To be able to assess the possibility of biased associations due to non-random missingness of the data, we used inverse probability weighting (IPW). This method has been recommended in epidemiological research over multiple imputation in situations where blocks of data are missing which is often the reason for attrition in ALSPAC where a variable is often missing due to non-participation in a clinic assessment visit (Weavers et al., 2021) In IPW complete cases are weighted by the inverse of their probability of being a complete case and involves conducting a response/non-response model in order to account for any bias in patterns of association due to missingness (Seaman & White, 2013).

For primary analyses, selective dropout was determined by comparing those participants who completed the HCL-32 questionnaire to those who dropped out, using logistic regression analyses. In line with previous publications based on this cohort (Marwaha et al., 2018; Morales-Muñoz et al., 2022, 2023) we used data from early time points in the ALSPAC dataset on child's sex and ethnicity, mother's socioeconomic status, maternal age, birthweight, preterm delivery and family adversity, to predict missingness in our analysis sample. The individuals associated with attrition at 21-23 years were more often males, and their mothers had higher scores on Family Adversity Index, they had lower socioeconomic status, had younger mother at birth (see Table S3).

Subsequently, we conducted IPW to account for those lost to follow-up. In accordance with previous research,(Morales-Muñoz et al., 2020, 2024) we used the variables associated with selective drop-out as the independent variables and fitted a logistic regression model (response vs. nonresponse as outcome) to determine weights for each individual using the inverse probability of response. Associations were not similar for the unweighted and weighted data, and thus we used the weighted data in all subsequent analysis.

For the secondary analyses, selective dropout was also determined by comparing those participants who completed the Stop Signal and N-Back questionnaire to those who dropped out, using two separate logistic regression analyses; one for participants who completed the Stop Signal and one for participants who completed the N-Back. The individuals associated with attrition at 24 years for completing the Stop Signal (see Table S4) and N-Back test (see Table S5) were more often males, and their mothers had higher scores on Family Adversity Index, they had lower socioeconomic status, had younger mother at birth.

*S3.5 Cross-tabulations for sparse cells/separation*

When the logistic regression models returned 0.00 odds ratios and confidence intervals for some classes, we conducted follow-up cross-tabulations between sleep trajectory classes and the binary hypomania variable to investigate potential sparse or empty cells (see Supplementary Tables S1–S2). Cross-tabulation of nighttime sleep duration trajectory classes and hypomanic symptom status confirmed that no participants in the persistently short sleep duration class (n = 16) met criteria for hypomanic symptoms (0 %). This complete separation explains the 0.00 odds ratio and 0–0 confidence interval reported for this class in Table 3. Given the small size but distinct developmental profile of this group, it was retained for completeness but excluded from inferential interpretation. The full cross-tabulation results are provided in Supplementary Table S1.

Cross-tabulation of the chronotype (MPS) classes indicated that the persistently later MPS class (n = 150) contained only one participant with hypomanic symptoms (0.7 %), leading to quasi-complete separation. This sparsity yielded an odds ratio of 0.00 with a 0–0 confidence interval in the weighted model. The class was nevertheless retained given its theoretical relevance in capturing a later-sleep phenotype during childhood. The full cross-tabulation results are presented in Supplementary Table S2.

### Supplementary Results

#### S4. Additional information on interaction models

*S4.1 Working memory models*

*Interaction between average short sleep duration in childhood and also experiencing hypomanic symptoms at 21-23 years on working memory at 24 years*

The adjusted model was significant, $F(9, 359) = 2.27$, $p = 0.017$, explaining 5.4% of the variance (adjusted $R^2 = 0.030$). Neither hypomanic symptoms (B = -0.72, SE = 0.51, 95% CI [-1.73, 0.29], p = 0.160) nor average short sleep duration (B = 0.08, SE = 0.10, 95% CI [–0.11, 0.28], p = 0.408) were significantly associated with n-back task performance at age 24. The interaction between average short sleep duration and hypomanic symptoms was not significant either (B = 0.56, SE = 0.82, 95% CI [–1.05, 2.17], p = 0.493).

*Interaction between persistently long sleep duration in childhood and also experiencing hypomanic symptoms at 21-23 years on working memory at 24 years*

The adjusted model was significant, $F(9, 1643) = 12.92$, $p < 0.001$, explaining 6.6% of the variance (adjusted $R^2 = 0.061$). Hypomanic symptoms were significantly associated with poorer n-back task performance (B = -0.41, SE = 0.16, 95% CI [-0.71, -0.10], p = 0.009). Persistently long sleep duration was not associated with n-back task performance (B = -0.07, SE = 0.07, 95% CI [-0.20, 0.07], p = 0.322). The interaction between persistently long sleep duration and hypomanic symptoms was not significant (B = 0.66, SE = 0.34, 95% CI [-0.01, 1.34], p = 0.053).

*Interaction between persistently early MPS in childhood and also experiencing hypomanic symptoms at 21-23 years on working memory at 24 years*

The adjusted model was significant, $F(9, 1448) = 10.48$, $p < 0.001$, explaining 6.1% of the variance (adjusted $R^2 = 0.055$). Hypomanic symptoms were significantly associated with poorer n-back task performance (B = −0.694, SE = 0.21, 95% CI [-1.11, -0.28], p = 0.001). Early MPS trajectory class was not associated with n-back task performance (B = -0.02, SE = 0.04, 95% CI [-0.11, 0.07], p = 0.650). The interaction between early MPS class and hypomanic symptoms was not significant (B = 0.71, SE = 0.39, 95% CI [–0.05, 1.48], p = 0.067).

*S4.2 Response inhibition models*

*Interaction between persistently early MPS in childhood and also experiencing hypomanic symptoms at 21-23 years on response inhibition at 24 years*

For Stop Signal Task performance, the adjusted model was significant, $F(9, 1526) = 14.25$, $p < 0.001$, explaining 7.8% of the variance (adjusted $R^2 = 0.072$). Hypomanic symptoms were significantly associated with higher SSRTs (B = 47.71, SE = 11.73, 95% CI [24.70, 70.71], $p <0.001$), suggesting worse response inhibition. Persistently early MPS was not associated with SSRT performance (B = 3.96, SE = 2.66, 95% CI [-1.25, 9.17], p = 0.136). The interaction between persistently early MPS and hypomanic symptoms was not significant (B = -26.27, SE = 27.77, 95% CI [-80.73, 28.20], p = 0.344).

*Interaction between average short sleep duration in childhood and also experiencing hypomanic symptoms at 21-23 years on response inhibition at 24 years*

The adjusted model was significant, $F(9,3012) = 40.59$, $p < 0.001$, explaining 10.8% of the variance (adjusted $R^2 = 0.106$). While hypomanic symptoms were not associated with SSRT performance (B = 15.08, SE = 9.31, 95% CI [-3.17, 33.33], p = 0.105), average short sleep duration was associated with higher SSRTs, suggesting worse response inhibition (B = 11.19, SE = 2.10, 95% CI [7.08, 15.30], $p <0.001$). The interaction between average short sleep duration and hypomanic symptoms was significant (B = 36.55, SE = 14.39, 95% CI [8.33, 64.78], p = 0.011), indicating that individuals with both risk factors performed significantly worse on SSRTs compared to those without both risk factors.

*Interaction between persistently long sleep duration in childhood and also experiencing hypomanic symptoms at 21-23 years on response inhibition at 24 years*

The overall model was significant, $F(9,1526) = 15.555$, $p < 0.001$, explaining 8.4% of the variance (adjusted $R^2 = 0.079$). Hypomanic symptoms were significantly associated with higher SSRTs (B = 55.98, SE = 11.30, 95% CI [33.81, 78.15], $p < 0.001$), suggesting worse response inhibition. Persistently long sleep duration was not associated with SSRT performance (B = -1.93, SE = 4.13, 95% CI [-10.03, 6.18], p = 0.641). The interaction between persistently long sleep duration and hypomanic symptoms was significant (B = -125.21, SE = 34.84, 95% CI [-193.56, -56.86], $p < 0.001$), indicating that participants with both persistently long sleep duration in childhood and hypomanic symptoms at 21–23 years demonstrated lower SSRTs, suggesting better response inhibition.

**Supplementary Tables**

**Table S1. Cross-tabulation of Night-time Sleep Duration Trajectory Classes and Hypomanic Symptoms**

| Night-time Sleep Duration Trajectory Class | Hypomanic Symptoms = No | Hypomanic Symptoms = Yes | Total N | Dichotomous HS variable at 21–23 years (% Yes) |
|---|---|---|---|---|
| Persistently average longer (Ref) | 993 | 16 | 1009 | 1.6 % |
| Persistently average shorter | 470 | 8 | 478 | 1.7 % |
| Persistently longer | 174 | 1 | 175 | 0.6 % |
| Persistently shorter | 16 | 0 | 16 | 0 % |
| Total | 1653 | 25 | 1678 | 1.5% |

*Note.* The persistently short sleep duration class contained no participants with hypomanic symptoms, resulting in complete separation; thus, odds ratios and confidence intervals could not be estimated (reported as 0.00). Of the 1,694 participants with classifiable hypomanic-symptom status, 1,678 had an assigned sleep-duration trajectory class. The 16 without an assigned class were all classified as not experiencing hypomanic symptoms.

**Table S2. Cross-tabulation of Chronotype (MPS) Trajectory Classes and Hypomanic Symptoms**

| Midpoint of Sleep Trajectory Classes | Hypomanic Symptoms = No | Hypomanic Symptoms = Yes | Total N | Dichotomous HS variable at 21–23 years (% Yes) |
|---|---|---|---|---|
| Persistently earlier MPS | 635 | 8 | 643 | 1.2 % |
| Persistently later MPS | 149 | 1 | 150 | 0.7 % |
| Persistently average earlier MPS (Ref) | 869 | 16 | 885 | 1.8 % |
| Total | 1653 | 25 | 1678 | 1.5 % |

*Note.* The persistent later MPS class contained only one participant with clinically significant hypomanic symptoms, leading to quasi-complete separation; odds ratios and confidence intervals could therefore not be reliably estimated and are reported as 0.00. Of the 1,694 participants with classifiable hypomanic-symptom status, 1,678 had an assigned MPS trajectory class. The 16 without an assigned class were the same participants omitted from Table S1 and were all classified as not experiencing hypomanic symptoms.

**Table S3. Differences in socio-demographic variables between non-participating and participating individuals in the study for hypomanic symptoms at age 21-23 years**

| | **Non-participating group in the study** | | **Participating group in the study** | | **Non-participating versus participating** | |
|---|---|---|---|---|---|---|
| | *Mean* | *SD* | *Mean* | *SD* | *OR (95% CI)* | *p* |
| Family Adversity Score | 4.54 | 4.39 | 3.46 | 3.67 | 0.96 (0.94-0.98) | <.001 |
| Mother's Socioeconomic Status | 52.22 | 13.53 | 56.45 | 13.45 | 1.01 (1.00-1.01) | 0.014 |
| Birthweight | 3.38 | 58.59 | 3.42 | 5.39 | 1.00 (1.00) | 0.418 |
| Maternal Age | 28.15 | 4.86 | 29.69 | 4.35 | 1.04 (1.02-1.06) | <.001 |
| | **Non-participating group in the study** | | **Participating group in the study** | | | |
| | *N* | *%* | *N* | *%* | | |
| Sex | | | | | 2.10 (1.79-2.46) | <.001 |
| Male/Female | 7082 / 6255 | 53.1 / 46.9 | 602 / 1091 | 35.6 / 64.6 | | |
| Ethnicity | | | | | 0.95 (0.62-1.46) | 0.815 |
| White/Other | 10011 / 554 | 94.8 / 5.2 | 1506 / 58 | 96.3 / 3.7 | | |
| Preterm Delivery | | | | | 0.76 (0.52-1.11) | 0.158 |
| Yes/No | 793 / 6663 | 10.6 / 89.4 | 946 / 63 | 93.8 / 6.2 | | |

*Note: Participating refers to individuals with HCL-32 questionnaire data at ages 21–23; non-participating refers to individuals without these data.*

**Table S4. Differences in socio-demographic variables between non-participating and participating individuals in the study for the Stop Signal test at age 24 years**

| | Non-participating group in the study | | Participating group in the study | | Non-participating versus participating | |
|---|---|---|---|---|---|---|
| | *Mean* | *SD* | *Mean* | *SD* | *OR (95% CI)* | *p* |
| Family Adversity Score | -74.67 | 1540.40 | 3.30 | 3.77 | 0.94 (0.93-0.96) | <0.001 |
| Mother's Socioeconomic Status | 51.34 | 13.27 | 57.02 | 13.65 | 1.02 (1.01-1.02) | <0.001 |
| Birthweight | 3374.2 | 593.4 | 3404.8 | 535.73 | 1.00 (1.00-1.00) | 0.726 |
| Maternal Age | 27.55 | 5.00 | 29.49 | 4.54 | 1.06 (1.04-1.07) | <0.001 |
| | **Non-participating group in the study** | | **Participating group in the study** | | | |
| | *N* | *%* | *N* | *%* | | |
| Sex | | | | | 2.11 (1.9-2.37) | <0.001 |
| Male/Female | 6401/5200 | 83.3 / 70.8 | 1283/2146 | 16.7 / 29.2 | | |
| Ethnicity | | | | | 0.79 (0.58-1.09) | 0.153 |
| White/Other | 8571/494 | 74.4 / 80.6 | 2949/119 | 25.6/ 19.4 | | |
| Preterm Delivery | | | | | 0.99 (0.76-1.28) | 0.922 |
| Yes/No | 704/5754 | 82.2/75.6 | 152/1856 | 17.8 / 24.4 | | |

*Note:* Participating refers to individuals with Stop Signal Task data at age 24; non-participating refers to individuals without these data.

**Table S5. Differences in socio-demographic variables between non-participating and participating subjects in the study for the N-Back test at age 24 years**

| | Non-participating group in the study | | Participating group in the study | | Non-participating versus participating | |
|---|---|---|---|---|---|---|
| | *Mean* | *SD* | *Mean* | *SD* | *OR (95% CI)* | *p* |
| Family Adversity Score | -64.30 | 1434.28 | 3.44 | 3.86 | 0.96 (0.95-0.98) | <0.001 |
| Mother's Socioeconomic Status | 52.30 | 13.63 | 55.62 | 12.99 | 1.01 (1.00-1.01) | 0.003 |
| Birthweight | 3378.8 | 586.72 | 3397.3 | 538.43 | 1.00 (1.00-1.00) | 0.631 |
| Maternal Age | 27.83 | 5.01 | 29.13 | 4.50 | 1.03 (1.01-1.05) | <0.001 |
| | **Non-participating group in the study** | | **Participating group in the study** | | | |
| | *N* | *%* | *N* | *%* | | |
| Sex | | | | | 2.21 (1.90-2.56) | <0.001 |
| Male/Female | 7045/6091 | 91.7 / 82.9 | 639/1255 | 8.3/17.1 | | |
| Ethnicity | | | | | 0.74 (0.49-1.11) | 0.148 |
| White/Other | 9891/555 | 85.9 / 90.5 | 1629/58 | 14.1/9.5 | | |
| Preterm Delivery | | | | | 1.10 (0.80-1.50) | 0.561 |
| Yes/No | 764/6580 | 89.3/86.5 | 92/1030 | 10.7/13.5 | | |

*Note:* Participating refers to individuals with n-back task data at age 24; non-participating refers to individuals without these data.

**Supplementary Figures**

**Figure S1.** Attrition of participants from the ALSPAC cohort

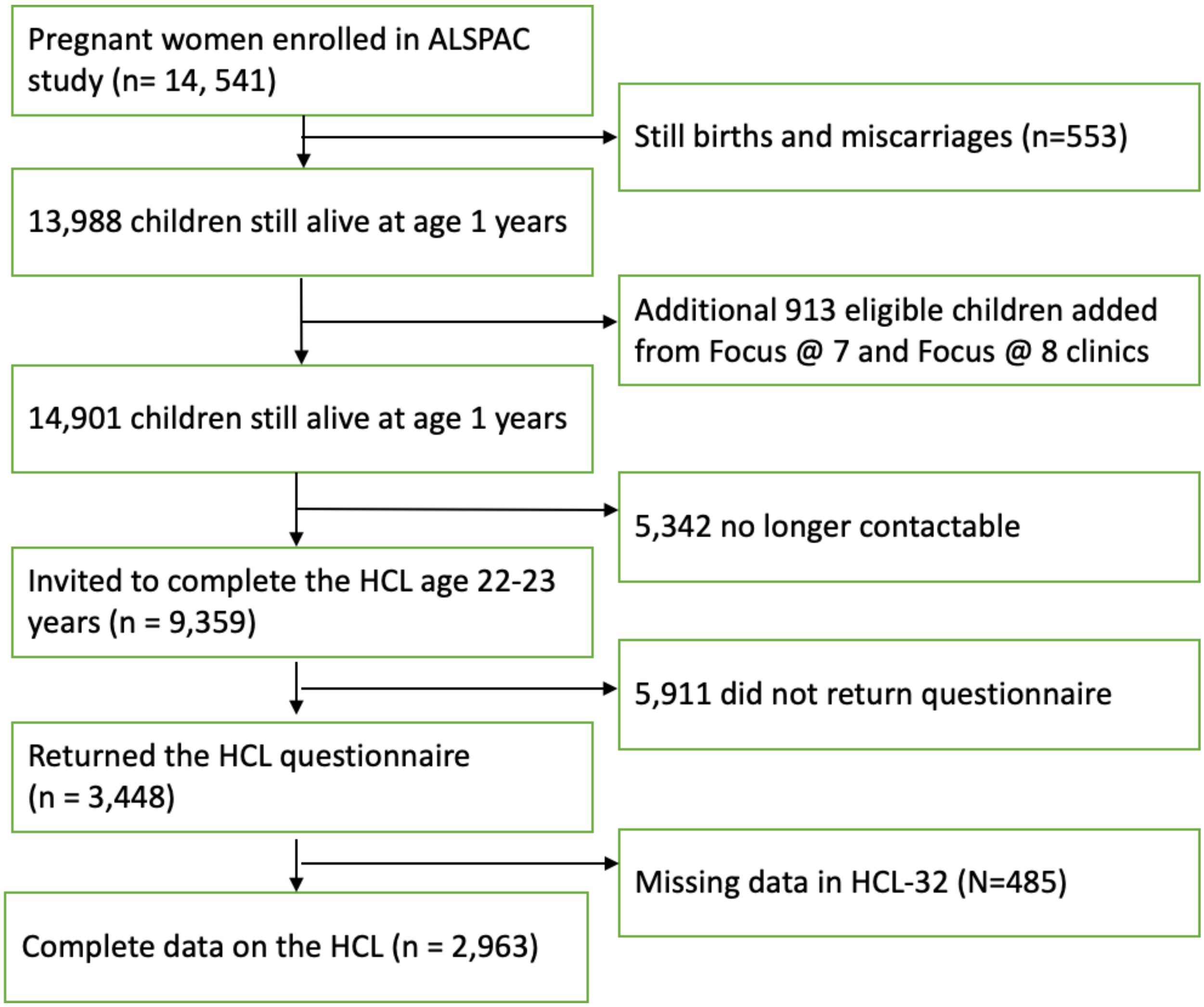

### Supplementary References


Angst, J., Azorin, J. M., Bowden, C. L., Perugi, G., Vieta, E., Gamma, A., & Young, A. H. (2011). Prevalence and characteristics of undiagnosed bipolar disorders in patients with a major depressive episode: The BRIDGE study. *Archives of General Psychiatry*, *68*(8), 791–799. https://doi.org/10.1001/archgenpsychiatry.2011.87

Bergink, V., Larsen, J. T., Hillegers, M. H. J., Dahl, S. K., Stevens, H., Mortensen, P. B., Petersen, L., & Munk-Olsen, T. (2016). Childhood adverse life events and parental psychopathology as risk factors for bipolar disorder. *Translational Psychiatry*, *6*(10), 2–7. https://doi.org/10.1038/TP.2016.201

Boyd, A., Golding, J., Macleod, J., Lawlor, D. A., Fraser, A., Henderson, J., Molloy, L., Ness, A., Ring, S., & Smith, G. D. (2013). Cohort profile: The 'Children of the 90s'-The index offspring of the avon longitudinal study of parents and children. *International Journal of Epidemiology*, *42*(1), 111–127. https://doi.org/10.1093/ije/dys064

Carvalho, A. F., Takwoingi, Y., Sales, P. M. G., Soczynska, J. K., Köhler, C. A., Freitas, T. H., Quevedo, J., Hyphantis, T. N., McIntyre, R. S., & Vieta, E. (2015). Screening for bipolar spectrum disorders: A comprehensive meta-analysis of accuracy studies. *Journal of Affective Disorders*, *172*, 337–346. https://doi.org/10.1016/j.jad.2014.10.024

Clemente, A. S., Diniz, B. S., Nicolato, R., Kapczinski, F. P., Soares, J. C., Firmo, J. O., & Castro-Costa, É. (2015). Bipolar disorder prevalence: A systematic review and meta-analysis of the literature. *Revista Brasileira de Psiquiatria*, *37*(2), 155–161. https://doi.org/10.1590/1516-4446-2012-1693

Court, H., Forty, L., Jones, L., Gordon-Smith, K., Jones, I., Craddock, N., & Smith, D. J. (2014). Improving the psychometric utility of the hypomania checklist (HCL-32): A Rasch analysis approach. *Journal of Affective Disorders*, *152–154*(1), 448–453. https://doi.org/10.1016/j.jad.2013.10.014

Dell'Osso, B., Cafaro, R., & Ketter, T. A. (2021). Has Bipolar Disorder become a predominantly female gender related condition? Analysis of recently published large sample studies. *International Journal of Bipolar Disorders*, *9*(1). https://doi.org/10.1186/s40345-020-00207-z

Forty, L., Smith, D., Jones, L., Jones, I., Caesar, S., Fraser, C., Gordon-Smith, K., & Craddock, N. (2009). Identifying hypomanic features in major depressive disorder using the hypomania checklist (HCL-32). *Journal of Affective Disorders*, *114*(1–3), 68–73. https://doi.org/10.1016/j.jad.2008.07.017

Fraser, A., Macdonald-wallis, C., Tilling, K., Boyd, A., Golding, J., Davey smith, G., Henderson, J., Macleod, J., Molloy, L., Ness, A., Ring, S., Nelson, S. M., & Lawlor, D. A. (2013). Cohort profile: The avon longitudinal study of parents and children: ALSPAC mothers cohort. *International Journal of Epidemiology*, *42*(1), 97–110. https://doi.org/10.1093/ije/dys066

Harris, P. A., Taylor, R., Thielke, R., Payne, J., Gonzalez, N., & Conde, J. G. (2009). Research electronic data capture (REDCap)-A metadata-driven methodology and workflow process for providing translational research informatics support. *Journal of Biomedical Informatics*, *42*(2), 377–381. https://doi.org/10.1016/j.jbi.2008.08.010

Ivars, K., Nelson, N., Theodorsson, A., Theodorsson, E., Ström, J. O., & Mörelius, E. (2015). Development of salivary cortisol circadian rhythm and reference intervals in full-term infants. *PLoS ONE*, *10*(6), 1–13. https://doi.org/10.1371/journal.pone.0129502

Jones, H. J., Gage, S. H., Heron, J., Hickman, M., Lewis, G., Munafò, M. R., & Zammit, S. (2018). Association of combined patterns of tobacco and cannabis use in adolescence with psychotic experiences. *JAMA Psychiatry*, *75*(3), 240–246. https://doi.org/10.1001/jamapsychiatry.2017.4271

Kajeepeta, S., Gelaye, B., Jackson, C. L., & Williams, M. A. (2015). Adverse childhood experiences are associated with adult sleep disorders: A systematic review. *Sleep Medicine*, *16*(3), 320–330. https://doi.org/10.1016/j.sleep.2014.12.013

Kantermann, T., Sung, H., & Burgess, H. J. (2015). Comparing the Morningness-Eveningness Questionnaire and Munich ChronoType Questionnaire to the dim light melatonin onset. *Journal of Biological Rhythms*, *30*(5), 449–453. https://doi.org/10.1177/0748730415597520

Kirchner, W. K. (1958). AGE DIFFERENCES IN SHORT-TERM RETENTION OF RAPIDLY CHANGING INFORMATION Minnesota Mining and Manufacturing Company. *Journal of Experimental Psychology*, *55*(4), 352–358.

Logan, G. D., Cowan, W. B., & Davis, K. A. (1984). On the ability to inhibit simple and choice reaction time responses: A model and a method. *Journal of Experimental Psychology: Human Perception and Performance*, *10*(2), 276–291. https://doi.org/10.1037/0096-1523.10.2.276

Marwaha, S., Winsper, C., Bebbington, P., & Smith, D. (2018). Cannabis use and hypomania in young people: A prospective analysis. *Schizophrenia Bulletin*, *44*(6), 1267–1274. https://doi.org/10.1093/schbul/sbx158

Meyer, T. D., Hammelstein, P., Nilsson, L. G., Skeppar, P., Adolfsson, R., & Angst, J. (2007). The Hypomania Checklist (HCL-32): its factorial structure and association to indices of impairment in German and Swedish nonclinical samples. *Comprehensive Psychiatry*, *48*(1), 79–87. https://doi.org/10.1016/j.comppsych.2006.07.001

Meyer, T. D., Schrader, J., Ridley, M., & Lex, C. (2014a). The Hypomania Checklist (HCL) - Systematic review of its properties to screen for bipolar disorders. *Comprehensive Psychiatry*, *55*(5), 1310–1321. https://doi.org/10.1016/j.comppsych.2014.03.002

Meyer, T. D., Schrader, J., Ridley, M., & Lex, C. (2014b). The Hypomania Checklist (HCL) - Systematic review of its properties to screen for bipolar disorders. *Comprehensive Psychiatry*, *55*(5), 1310–1321. https://doi.org/10.1016/j.comppsych.2014.03.002

Morales-Muñoz, I., Broome, M. R., & Marwaha, S. (2020). Association of Parent-Reported Sleep Problems in Early Childhood with Psychotic and Borderline Personality Disorder Symptoms in Adolescence. *JAMA Psychiatry*, *77*(12), 1256–1265. https://doi.org/10.1001/jamapsychiatry.2020.1875

Morales-Muñoz, I., Mallikarjun, P. K., Chandan, J. S., Thayakaran, R., Upthegrove, R., & Marwaha, S. (2023). Impact of anxiety and depression across childhood and adolescence on adverse outcomes in young adulthood: a UK birth cohort study. *British Journal of Psychiatry*, *45*(4), 212–220. https://doi.org/10.1192/bjp.2023.23

Morales-Muñoz, I., Marwaha, S., Upthegrove, R., & Cropley, V. (2024). Role of Inflammation in Short Sleep Duration Across Childhood and Psychosis in Young Adulthood. *JAMA Psychiatry*, 1–9. https://doi.org/10.1001/jamapsychiatry.2024.0796

Morales-Muñoz, I., Palmer, E. R., Marwaha, S., Mallikarjun, P. K., & Upthegrove, R. (2022). Persistent Childhood and Adolescent Anxiety and Risk for Psychosis: A Longitudinal Birth Cohort Study. *Biological Psychiatry*, *92*(4), 275–282. https://doi.org/10.1016/j.biopsych.2021.12.003

Morgan, G. B. (2015). Mixed Mode Latent Class Analysis: An Examination of Fit Index Performance for Classification. *Structural Equation Modeling*, *22*(1), 76–86. https://doi.org/10.1080/10705511.2014.935751

Nosarti, C., Reichenberg, A., Murray, R. M., Cnattingius, S., Lambe, M. P., Yin, L., MacCabe, J., Rifkin, L., & Hultman, C. M. (2012). Preterm birth and psychiatric disorders in young adult life. *Archives of General Psychiatry*, *69*(6), 1–8. https://doi.org/10.1001/archgenpsychiatry.2011.1374

Paksarian, D., Cui, L., Angst, J., Ajdacic-Gross, V., Rössler, W., & Merikangas, K. R. (2016). Latent trajectories of common mental health disorder risk across 3 decades of adulthood in a population-based cohort. *JAMA Psychiatry*, *73*(10), 1023–1031. https://doi.org/10.1001/jamapsychiatry.2016.1921

Palmier-Claus, J. E., Berry, K., Bucci, S., Mansell, W., & Varese, F. (2016). Relationship between childhood adversity and bipolar affective disorder: Systematic review and meta-analysis. *British Journal of Psychiatry*, *209*(6), 454–459. https://doi.org/10.1192/bjp.bp.115.179655

Polga, N., Macul Ferreira de Barros, P., Farhat, L. C., de Almeida, K. M., Bloch, M. H., & Lafer, B. (2022). Parental age and the risk of bipolar disorder in the offspring: A systematic review and meta-analysis. *Acta Psychiatrica Scandinavica*, *145*(6), 568–577. https://doi.org/10.1111/acps.13418

Rivkees, S. A. (2003). Developing circadian rhythmicity in infants. *Pediatric Endocrinology Reviews*, *1*(1), 38–45.

Seaman, S. R., & White, I. R. (2013). Review of inverse probability weighting for dealing with missing data. *Statistical Methods in Medical Research*, *22*(3), 278–295. https://doi.org/10.1177/0962280210395740

Simpkin, C. T., Jenni, O. G., Carskadon, M. A., Wright, K. P., Akacem, L. D., Garlo, K. G., & Lebourgeois, M. K. (2014). Chronotype is associated with the timing of the circadian clock and sleep in toddlers. *Journal of Sleep Research*, *23*(4), 397–405. https://doi.org/10.1111/jsr.12142

Smith, D. J., Anderson, J., Zammit, S., Meyer, T. D., Pell, J. P., & Mackay, D. (2015). Childhood IQ and risk of bipolar disorder in adulthood: prospective birth cohort study. *BJPsych Open*, *1*(1), 74–80. https://doi.org/10.1192/bjpo.bp.115.000455

Tofighi, D., & Enders, C. K. (2008). Identifying the correct number of classes in growth mixture models. *Advances in Latent Variable Mixture Models*, 317–341.

Weavers, B., Heron, J., Thapar, A. K., Stephens, A., Lennon, J., Bevan Jones, R., Eyre, O., Anney, R. J., Collishaw, S., Thapar, A., & Rice, F. (2021). The antecedents and outcomes of persistent and remitting adolescent depressive symptom trajectories: a longitudinal, population-based English study. *The Lancet Psychiatry*, *8*(12), 1053–1061. https://doi.org/10.1016/S2215-0366(21)00281-9

Winsper, C., Hall, J., Strauss, V. Y., & Wolke, D. (2017). Aetiological pathways to Borderline Personality Disorder symptoms in early adolescence: Childhood dysregulated behaviour, maladaptive parenting and bully victimisation. *Borderline Personality Disorder and Emotion Dysregulation*, *4*(1), 1–10. https://doi.org/10.1186/s40479-017-0060-x

Winsper, C., Tang, N. K. Y., Marwaha, S., Lereya, S. T., Gibbs, M., Thompson, A., & Singh, S. P. (2017). The sleep phenotype of Borderline Personality Disorder: A systematic review and meta-analysis. *Neuroscience and Biobehavioral Reviews*, *73*, 48–67. https://doi.org/10.1016/j.neubiorev.2016.12.008

Winsper, C., Wolke, D., Scott, J., Sharp, C., Thompson, A., & Marwaha, S. (2020). Psychopathological outcomes of adolescent borderline personality disorder symptoms. *Australian and New Zealand Journal of Psychiatry*, *54*(3), 308–317. https://doi.org/10.1177/0004867419882494

Wolke, D., Schreier, A., Zanarini, M. C., & Winsper, C. (2012). Bullied by peers in childhood and borderline personality symptoms at 11 years of age: A prospective study. *Journal of Child Psychology and Psychiatry and Allied Disciplines*, *53*(8), 846–855. https://doi.org/10.1111/j.1469-7610.2012.02542.x

Zanarini, M. C. (2003). Zanarini Rating Scale for Borderline Personality Disorder (ZAN-BPD): A continuous measure of DSM-IV borderline psychopathology. *Journal of Personality Disorders*, *17*(3), 233–242. https://doi.org/10.1521/pedi.17.3.233.22147

Zanarini, M. C., & Frankenburg, F. R. (2001). Attainment and maintenance of reliability of axis I and II disorders over the course of a longitudinal study. *Comprehensive Psychiatry*, *42*(5), 369–374. https://doi.org/10.1053/comp.2001.24556
